# Positive Affect as a Developmental Mediator of Early Adversity and Internalizing Psychopathology

**DOI:** 10.1101/2025.08.01.25332831

**Authors:** Jamie L. Hanson, Dorthea J Adkins, Isabella Kahhale, Sriparna Sen

## Abstract

**Background:** Early life adversities (ELAs) including experiences such as abuse, neglect, and household dysfunction are strongly linked to psychopathology; yet, the developmental pathways connecting ELA to externalizing and internalizing psychopathology remain unclear. While most research has focused on threat and negative affect, positive emotions may represent a critical but understudied mechanism linking ELA to mental health outcomes.

**Methods:** Using data from the Adolescent Brain Cognitive Development (ABCD) study, we examined positive affect trajectories across six timepoints spanning childhood through adolescence (ages 9-10 to 12-13). We employed person-centered trajectory-based clustering to identify distinct patterns in positive affect – independent of ELA exposure – followed by multinomial logistic regression to examine associations between cumulative ELA exposure and trajectory membership. Mediation analyses tested whether positive affect trajectories explained links between ELA and psychopathology outcomes.

**Results:** Four distinct positive affect trajectories emerged: High-Stable, Declining, Persistently Low, and Volatile (N=7,457). Higher ELA scores significantly predicted membership in all non-high-stable trajectories, with the strongest association existing for the Persistently Low group (β = 0.321, p < 0.001). Mediation analyses revealed that Persistently Low trajectory group membership significantly mediated the relationship between ELA and internalizing problems (indirect effect = 0.030, 95% CI [0.012, 0.056], p = 0.007), but not externalizing problems (N= 3,927).

**Conclusion:** This study demonstrates that ELA shapes positive affect development through distinct, heterogeneous pathways rather than uniform effects, with persistently low positive affect representing a specific mechanism linking early adversity to later depression and anxiety. Findings suggest that targeting positive emotional experiences may be a promising intervention strategy for youth exposed to ELA.

---

Experiences of early life adversity (ELA) are unfortunately common and can profoundly shape youth mental health and development (Bhutta et al., 2023; Gilgoff et al., 2020). Millions of children both nation-wide and globally face ELAs such as physical abuse and neglect (Madigan et al., 2023; Sacks & Murphey, 2018); such adversities are linked to a host of developmental challenges including difficulties in social-emotional skills and increases in different forms of psychopathology (Doyle & Cicchetti, 2017). Particularly notable is the connection between ELAs and the emergence and development of internalizing symptoms in children and adolescents (Green et al., 2010). Meta-analyses examining multiple forms of ELA find that youth with a history of ELA are not only more likely to develop depression and anxiety, but also have more severe presentations of these conditions, poorer treatment prognosis, and lower remission rates compared to youth without ELA (Espejo et al., 2007; Humphreys et al., 2020; LeMoult et al., 2020; Mandelli et al., 2015; Nanni et al., 2012; Tyrka et al., 2013; Wiersma et al., 2009; Williams et al., 2016). Despite such findings, the pathways leading from ELA to depression remain unclear. Clarifying the developmental mechanisms linking ELA to depression is essential for designing targeted, developmentally-informed interventions that improve outcomes for youth exposed to adversity.

The majority of past investigations on ELA and factors contributing to poor mental health have focused on threat and negative affect, finding that altered threat- and emotion processing of negative affect following ELA is associated with subsequent psychopathology (Glaser et al., 2006; McLaughlin et al., 2009; Pollak & Kistler, 2002). However, scarce work has explored potential alterations in *positive* affect (or lack thereof) after ELA. Positive affect can be conceptualized as emotions or feelings reflecting enthusiasm, energy, and pleasurable engagement with the environment such as happiness, joy, excitement and contentment (Alexander et al., 2021; Clark et al., 1989; Watson, Clark, & Tellegen, 1988). Importantly, positive and negative affect represent largely independent dimensions of emotional experience rather than opposite poles of a single continuum (Watson, Clark, & Carey, 1988; Watson & Tellegen, 1985). Factor analytic studies underscore that affective systems can be dissociated, i.e., individuals may experience high levels of both positive and negative affect simultaneously, or show deficits in one domain while the other remains intact (Cacioppo et al., 1999; Diener & Emmons, 1984). This independence suggests that the mechanisms linking ELA to altered positive affect may be distinct from those involving negative emotional processing. The limited focus on positive affect in the literature on ELA and depression and anxiety is, however, notable given that self-reported low positive affect, and lower reactivity to positive stimuli, have been connected to depression, and to a lesser extent anxiety (Bylsma et al., 2008; Sequeira et al., 2022; Trøstheim et al., 2020). For example, individuals with low positive affect are 2-3 times more likely to develop later depression (Rackoff & Newman, 2020; Wood & Joseph, 2010). In addition, low positive affect has been found to relate to anxiety disorders, with consistent connections found for social anxiety disorders (Kashdan, 2007; Naragon-Gainey et al., 2009). Thinking more broadly, positive affect may be protective against other risk factors for depression and anxiety, as higher levels of these emotions can enhance the link between multiple resilience factors for depression such as social support, optimism, coping skills, and capacity to recover from negative events (Folkman, 2008; Khazanov & Ruscio, 2016; Southwick et al., 2005). Understanding the ways in which ELA may undermine positive affect is therefore essential to clarifying the role of positive affect in the developmental pathways to depression and anxiety.

ELA could negatively impact the development and expression of positive affect in several ways. First, ELAs could disrupt child-caregiver attachment, affecting how children interpret social interactions and relationships (Cyr et al., 2010). Insecure or disorganized attachment may influence a child’s ability to experience, recognize, and benefit from positive emotions and social connections by diminishing opportunities to learn normative social-emotional patterns (Palacios-Barrios et al., 2024). Second, exposure to abuse, neglect and other ELAs has been linked to dysregulated neuroendocrine and cortisol responsivity (Bunea et al., 2017; Shirtcliff et al., 2024); these alterations could influence behavioral responses to stress and cascade to emotion dysregulation. Finally, ELAs may influence the development of brain regions that are critical for learning, memory, cognition, and behavioral control (Gorka et al., 2014; Hanson et al., 2010, 2012; Nweze, Banaschewski, et al., 2023; Nweze, Ezenwa, et al., 2023). Taken together, these alterations to attachment, stress responsivity, cognition and neurobiology could compromise abilities to experience, express, and draw meaning from positive emotions.

Consistent with these mechanisms, multiple studies have found that childhood maltreatment – one form of ELA – is associated with lower positive affect and emotions in adults (DePierro et al., 2018; Etter et al., 2013; Kuzminskaite et al., 2024; Myroniuk et al., 2024; Turiano et al., 2017; Xiang et al., 2021). In child and adolescent samples, research in this area focused on positive affect has been limited. In young children (<4 years of age), laboratory tasks with caregivers designed to evoke positive affect or positive behaviors (e.g., cooperation and constructive play) have found lower levels of positive affect or behavior in children with high levels of ELA (Dadds et al., 2003; Egeland et al., 1983; Egeland & Sroufe, 1981; Robinson et al., 2009). Additional support for the hypothesis that ELAs may impact positive affect comes from meta-analyses finding that ELA leads to lower self-report on psychosocial constructs related to positive affect such as hope, sense of purpose, and self-concept (Hill et al., 2018; Melamed et al., 2024; Yarcheski & Mahon, 2016; Zhang et al., 2023). Further, children who suffer ELAs often struggle with understanding and recognizing positive emotions (Bick et al., 2017; Caouette et al., 2023; Diaconu et al., 2023; Fries & Pollak, 2004; Koizumi & Takagishi, 2014; Pears & Fisher, 2005), or forecasting situations that might lead to positive emotional outcomes (Perlman et al., 2008). Finally, ELAs also appear to impact behavioral responses to positive feedback and reward cues (Oltean et al., 2023) in adolescents (Hanson et al., 2017; Yazgan et al., 2021) and adults (Dillon et al., 2009). These findings collectively support the view that ELA undermines the capacity to experience, recognize, and respond to positive emotions.

While these past studies have provided important foundational insights, several key limitations exist that the current study seeks to overcome. First, emotional development across childhood and adolescence is dynamic, with youth learning how to better express, understand, and regulate their feelings throughout this time (Bailen et al., 2019). Research has found decreases in positive emotions during the transition from childhood to adolescence (Larson et al., 2002; Reitsema et al., 2022), necessitating a longitudinal understanding of these emotions’ trajectories. Second, scarce research on child and adolescent samples have measured positive affect *directly*, instead using observational tasks or constructs related to positive affect (e.g., positive feedback processing; reward learning). Measuring positive affect via self-report could be advantageous given that this emotion is a subjective, internal experience. Further, past work in *adult* samples that *have* directly measured positive affect has typically been cross-sectional and relies on retrospective reports of ELA, therefore introducing recall bias and prohibiting an understanding of how differences in positive affect may change over time in relation to ELA. Past research has also not adequately considered behavioral heterogeneity in how positive affect may be altered following ELA exposure (Gee, 2021; Hanson et al., 2024). Group-level analyses that examine mean differences may mask important subtypes of positive affect responsivity after adversity. Without person-centered approaches that can identify distinct patterns of positive affect functioning (Linden & Hönekopp, 2021), our ability to understand individual risk and resilience factors remains limited, preventing us from distinguishing youth with early positive affect declines from those with delayed onset deficits, or from those who show late-stage growth versus chronically low positive affect. Finally, previous projects have not typically connected differences in positive affect to symptoms of depression, anxiety or other forms of psychopathology.

Motivated by these limitations, we leveraged the Adolescent Brain Cognitive Development (ABCD) Study® to investigate the longitudinal impact of ELA on positive affect and psychopathology during childhood and adolescence. We hypothesized that youth who had experienced greater levels of ELA would show altered positive affect trajectories compared to youth with lower adversity exposure. We believed that, upon examining these person-centered positive affect trajectories, youth exposed to ELA would show early and consistent declines in positive affect, while others might exhibit declines later in development. Finally, we planned to examine relations between ELA, positive affect trajectories, and symptoms of psychopathology. We were interested in examining both *internalizing* (e.g., Depression; Anxiety) and *Externalizing* (e.g., Aggressive Behavior, Conduct Problems) symptoms. This would allow us to understand if positive affect was a distinct mechanism for internalizing issues, rather than a general psychopathology risk factor. We predicted that differences in positive affect trajectories would account for the relation between cumulative ELA exposure and *internalizing* psychopathology symptoms, underscoring the potential role of positive affect in linking early adversity to later mental health outcomes.

## Method

### Participants and Procedures

This work used data from the ABCD^®^ study, an ongoing longitudinal project that aims to understand how various factors influence youth development. Data was derived from the ABCD 5.1 release and included a baseline visit and multiple follow-up visits across the 4 subsequent years (see (Volkow et al., 2018) and the ABCD study website, https://abcdstudy.org/, for further details).

The ABCD study sampled individuals across 21 sites in the United States to reflect the nation’s diverse socio-demographic population. Researchers first created a catchment area which included public, private, and charter schools within 50 miles of a research site. Schools were then coded based on a series of factors (e.g. ethnic composition). The study used stratified sampling to randomly select schools from the catchment areas by presenting caregivers with recruitment materials. A total of 11,878 children were assessed at baseline (Karcher & Barch, 2021). Baseline data was collected in 2018 from a cohort of 9- to 10-year-old children and their caregivers.

### Ethical Information

Each ABCD recruitment site obtained full assent and consent from the children and their parent(s)/legal guardian(s), respectively in accordance with local Institutional Reviews Boards. Our work was reviewed by the University of Pittsburgh’s Institutional Review Board (ID: STUDY21020159). It was determined that our activity was not research involving human subjects as defined by U.S. Department of Health and Human Services and Food and Drug Administration regulations on 02/25/2021.

### Early Life Adversity (ELA)

ELA measures were identified from ABCD documentation and literature, reflecting categories from the commonly used Adverse Childhood Experiences (ACEs) scale (Felitti et al., 1998). A standardized cumulative risk score was derived from youth and caregiver reports across multiple baseline scales, as cumulative approaches have greater predictive power and overcome limitations of dimension-specific methods, including ill-defined boundaries, co-occurring adversities, and lack of specificity in effects (for review and discussion, see (Smith & Pollak, 2021). Assessing ELA during childhood also avoids retrospective recall biases inherent to adult self-report (Colman et al., 2016). Items were drawn from eight ABCD scales including the Kiddie Schedule for Affective Disorders and Schizophrenia (K-SADS), Family History Assessment, Neighborhood Safety, Parent Demographics, Family Environment Scale, Parental Monitoring, Children’s Report of Parent Behavior, and Parent Adult Self Report. The methodology involved averaging items within each scale, z-scoring responses at the scale level, standardizing scale averages across participants, and then averaging all z-scores to generate a final cumulative risk score for each participant. Inclusion required completion of at least 75% of ELA scales. The mean for this standardized composite was -0.09 (Standard Deviation=0.90) with a range from -1.09 to 12.61 (also see Figure S1). Additional information about this measure is detailed in our *Appendix S1 and Table S1*.

### Positive Affect

Positive affect describes pleasant emotions that arise when we feel engaged and satisfied with our environment, including feelings like happiness, joy, excitement, enthusiasm, and contentment (Alexander et al., 2021). The Positive Affect Scale from the NIH Toolbox Battery (Salsman et al., 2013) was administered at different time points, but not at the baseline visit (i.e., after 6, 12, 18, 30, 36, and 42 months from the baseline visit), to evaluate positive emotions and affective well-being in the past week. Respondents were provided with 9 positive affect items (e.g., “I felt calm”, “I felt delighted”) and indicated on a 3-point Likert scale (1 = Not true; 3 = Very true) how much they felt that way in the last week. The scale showed high internal validity in past publications (Cronbach’s alpha = .96). This measure was initially collected on N=11,734 participants; the current study examined participants with six waves of data (from follow-ups at 6-months, 1-year, 18-months, 30-months, 3-year, and 42-months).

### Psychopathology

Symptoms of psychopathology were measured via the Child Behavior Checklist (CBCL). The CBCL is a widely used parent-report measure that assesses youth behavioral problems and psychopathology occurring in the past 6 months on a 3-point scale (Achenbach, 2011). In the ABCD study, caregivers (primarily mothers) completed the CBCL during in-person visits. The CBCL generates multiple scales that empirically capture different behavioral and developmental problems. The current study focused on the *Total Internalizing Score* (e.g., Anxious/Depressed, Withdrawn/Depressed) and *Total Externalizing Score* (e.g., Aggressive Behavior, Conduct Problems) scores. Both internalizing and externalizing scores represent composite measures created by summing items across their respective subscales, with higher scores indicating more severe psychopathology and behavioral problems. We used raw scores from the baseline and the year 4 follow-up visits. Focusing on both internalizing and externalizing outcomes would allow us to speak to whether lower positive affect was a specific risk for one form of psychopathology or rather a general psychopathology risk factor.

### Covariates

The following variables assessed at baseline were included as covariates in sensitivity models: participant sex assigned at birth (0 = male, 1 = female), race/ethnicity (1 = White, 2= Black, 3 = Hispanic, 4 = Asian, 5 = Other), age in months, and recruitment site (dummy-coded). Additional sensitivity models controlled for socioeconomic status, as measured by household income. In line with past work (Hair et al., 2015; Hanson et al., 2011, 2013), the income variable was transformed by taking the midpoint of each income category and log-transforming this value to normalize the highly skewed distribution and ensure robust statistical modeling.

### Statistical Modeling

Data analysis consisted of three components: 1) person-center trajectory modeling of positive affect using time-series analyses; 2) multinomial regression models; and 3) mediation modeling combining multinomial and ordinary least squares regressions.

#### Person-centered trajectory modeling of positive affect

We cleaned and processed the longitudinal positive affect data by recoding invalid responses (e.g., 777, 999) as missing values and filtering out participants with four or more missing positive affect items. Only participants with complete data across all six waves were retained for analysis. Trajectory modeling using the R-package *traj* (Sylvestre et al., 2025; Sylvestre & Vatnik, 2014) involved calculating nineteen time-series indices assessing different aspects of longitudinal change patterns in individuals (Leffondré et al., 2004) including elementary measures of change (e.g., maximum, range, mean, standard deviation), linear modeling parameters (intercept, slope, R²), measures of nonlinearity and inconsistency (e.g., curve length, rate of intersection with mean), measures sensitive to nonmonotonicity and short-term fluctuations (first and second derivative statistics), and measures contrasting early versus later change. These measures are described in *Appendix S2* and noted in Table S2. Outliers were identified and capped (not removed) using a probability-based approach is a method conceptually similar to winsorizing (Nishiyama, 2018). Extreme values were defined as a threshold corresponding to a 0.3% probability and replaced with less extreme values allowing us to retain all observations while limiting the influence of extreme outliers on trajectory estimation. Next, highly correlated time-series indices (>0.95) were removed to reduce redundancy. The remaining indices underwent principal component analysis to identify a subset that captured the most important trajectory features, which were subsequently used for k-medoids clustering with 5,000 bootstrap samples and a maximum of 1,000 iterations. This analysis identified four distinct positive affect trajectory patterns. To determine the optimal number of clusters, we employed the Calinski-Harabasz criterion, which evaluates cluster quality by comparing the ratio of between-cluster to within-cluster variance (Caliński & Harabasz, 1974). Higher values indicate separation between trajectory groups while maintaining within-cluster homogeneity. This person-centered modeling was completed only for positive affect and was done independently of ELA and any other variables.

#### Multinomial regression

Models examined associations between cumulative ELA exposure and positive affect trajectory membership, controlling for relevant covariates (e.g., age, sex, race/ethnicity, socioeconomic status). These models estimated the likelihood of belonging to each trajectory cluster based on adversity exposure levels, with one cluster serving as the reference group (High-Stability Positive Affect). Multinomial logistic regression was selected because the trajectory clusters represent a categorical outcome variable with more than two unordered categories.

#### Mediation

To test whether positive affect trajectory clusters mediated the association between cumulative ELA exposure and psychopathology outcomes, we employed an approach that combined multinomial logistic regression (for the categorical mediator of trajectory cluster membership) with ordinary least squares regression (for continuous psychopathology outcomes). We decomposed the total effect of ELA on psychopathology into direct effects (ELA → psychopathology) and indirect effects (ELA → trajectory clusters → psychopathology), accounting for the categorical nature of the mediator variable. Mediation models using alternative formulations of internalizing subscales are detailed in *Appendix S3, Figure S2, and Table S3*.

#### Data loss

While ABCD collected data on over 11,000 participants, we had smaller analytic samples for our multinomial regression and mediation models. One major source of data loss was missing positive affect measures from the six available assessments, as N=4,277 participants did not have complete data on this questionnaire over tie. This led to a final analytic N=7,457 for our multinomial regression models examining associations between cumulative ELA exposure and positive affect trajectory cluster membership. With the mediation analyses, the analytic N=3,927, with N = 3,530 participants being excluded for missing data from the CBCL at the 4-year follow-up of the ABCD study. Our supplement contains flow charts noting where participant and how many participants were lost for different analyses (Figure S3), as well as missing completely at random (MCAR) analyses (also see *Appendix S4-S5)*.

#### Artificial Intelligence Generated Content

Claude Sonnett 4.5 was used in October of 2025, to update the R syntax that was used to generate tables and graphs, specifically those in *Appendix S3*. The first author takes responsibility for the integrity of the content generated.

## Results

Sociodemographic characteristics of the final sample are presented in **Table 1**.

**Table 1.** Sociodemographic Characteristics of Study Sample.

| Characteristic | Overall Sample<sup>1</sup> |
| --- | --- |
| Age at Baseline (Months) | 119.26 (7.47) |
| Sex |  |
| Male | 3,638 (52.2%) |
| Female | 3,326 (47.8%) |
| Race/Ethnicity |  |
| White | 4,200 (60.3%) |
| Hispanic | 1,279 (18.4%) |
| Black | 655 (9.4%) |
| Other/Mixed | 688 (9.9%) |
| Asian | 142 (2.0%) |
| Household Income |  |
| < \$25K | 304 (4.4%) |
| \$25K-\$49K | 736 (10.6%) |
| \$50K-\$99K | 2,617 (37.6%) |
| \$100K+ | 3,307 (47.5%) |

<sup>1</sup>Mean (SD); n (%)

### Person-centered trajectory modeling of positive affect

Using our non-parametric trajectory-based clustering approach, participants with complete follow-up data (N = 7,.457) were classified into 4 distinct clusters based on their trajectory of positive affect from ABCD time points 1 to 6 (**Figure 1A)**. Calinski-Harabasz values peaked at k=4 clusters (index = 0.717; **Figure 1B**). Notably, gap statistics were similar across different k values (range: 24.265-24.266) suggesting robust clustering structure, but with k=4 showing optimal separation (Calinski-Harabasz values=0.717). These groups were: 1) High and Stable (N = 2,055; 27.6%), which represented the largest subgroup and showed high positive affect, without significant declines over development; 2) Declining (N = 1,846; 24.8%), which had initially high levels of positive affect, but then showed significant decline during early adolescence; 3) Persistently Low (N = 1,817; 24.4%), which was characterized by consistently lower positive affect levels through childhood and adolescence; and 4) Volatile (N = 1,739; 23.3%), where positive affect was more variable and unstable across development. Demographics for each subgroup are noted in **Table 2**.

**Figure 1.**
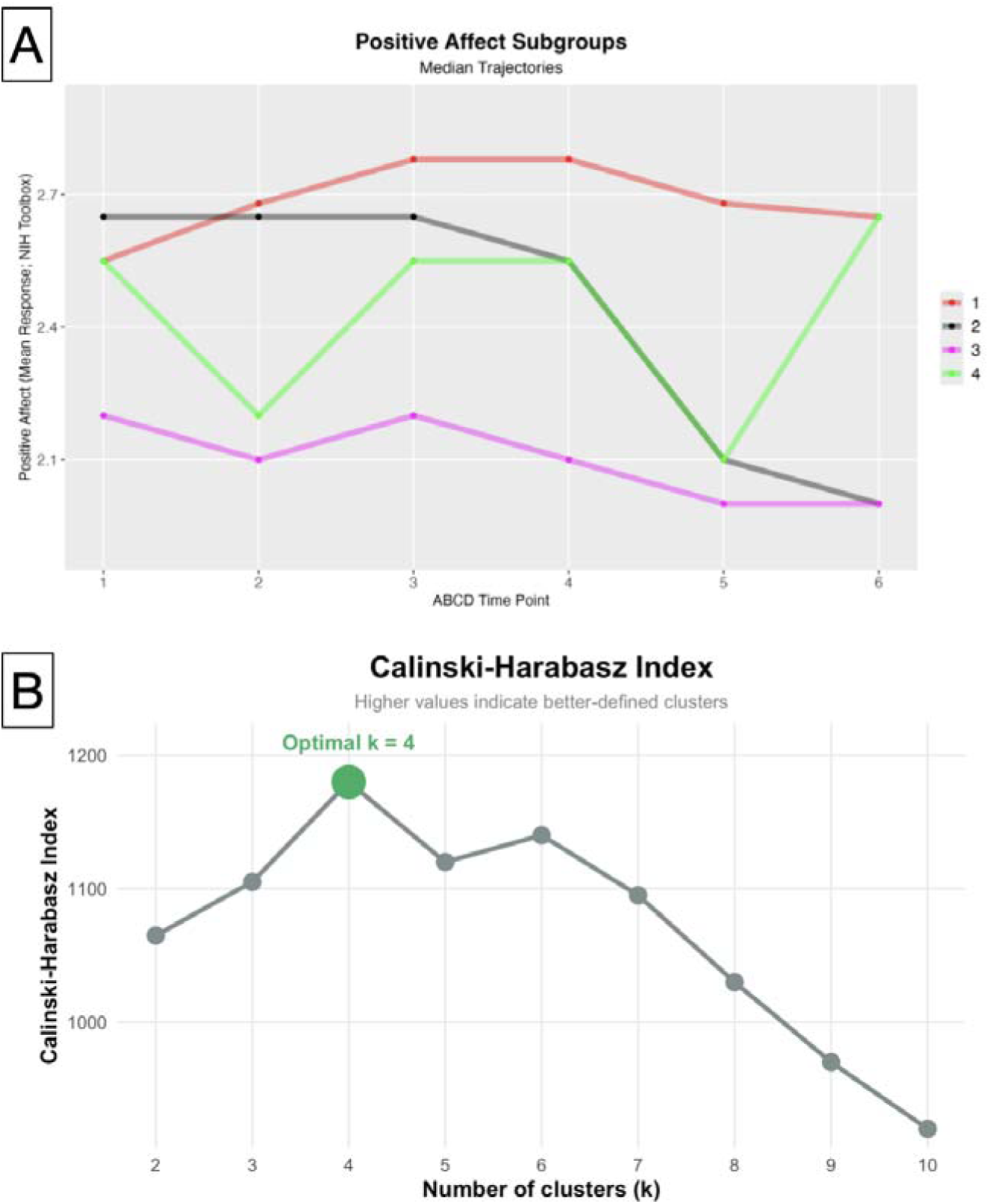
**Caption**: (A) Median positive affect trajectories across six ABCD study time points for four identified subgroups. Trajectories show positive affect mean response scores from the NIH Toolbox across developmental time points. Group 1 (red) shows high levels and primarily stability throughout development, Group 2 (black) demonstrates high levels initially, but then sharp declines in adolescence, Group 3 (magenta) exhibits persistently low levels, and Group 4 (green) displays a volatile pattern with significant fluctuations over time. (B) Calinski-Harabasz Index values across different clustering solutions (k = 2-10). The optimal solution of four clusters was identified at k = 4 (peak value = 1,180), indicating the most distinct and well-separated trajectory subgroups.

**Table 2.**
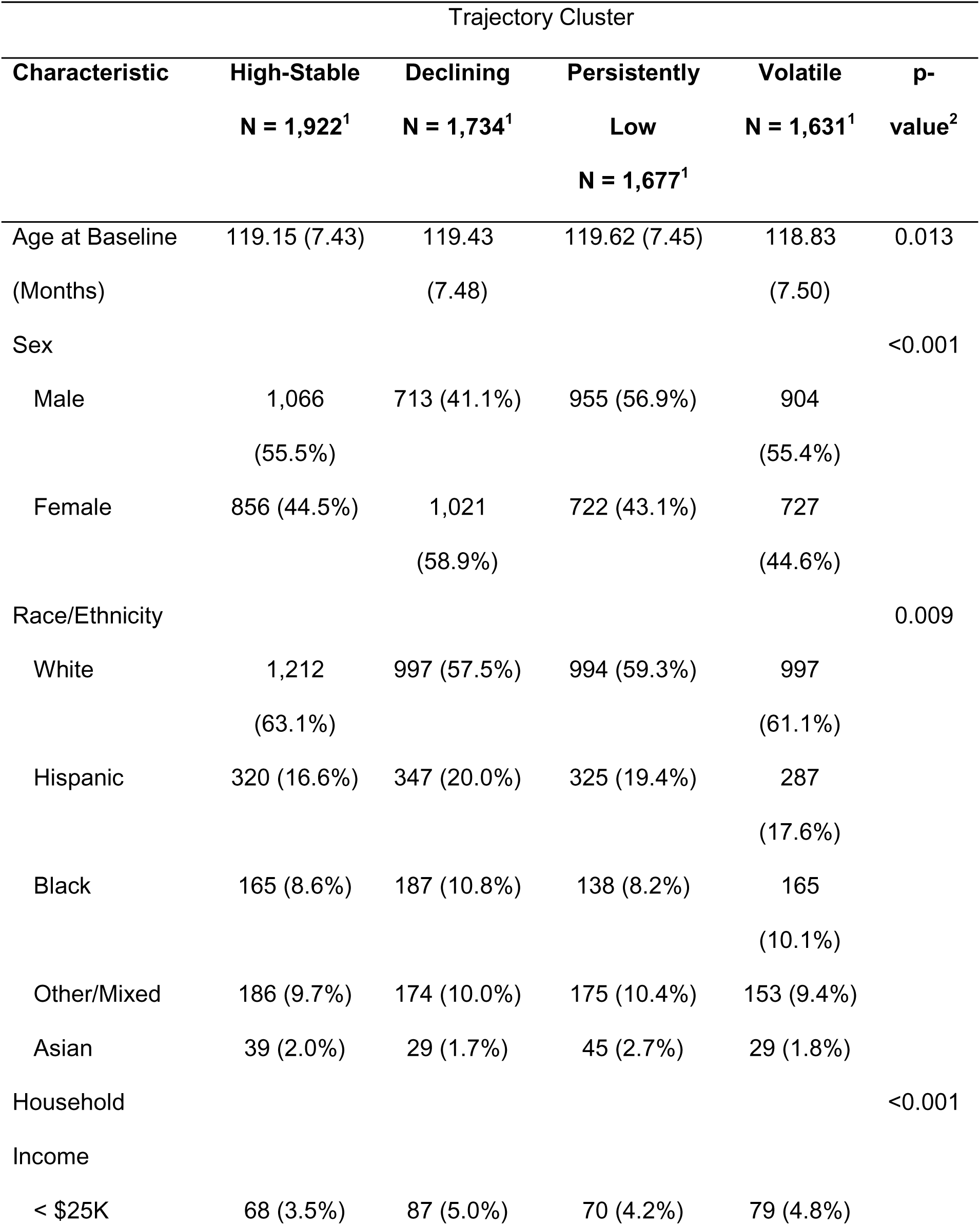
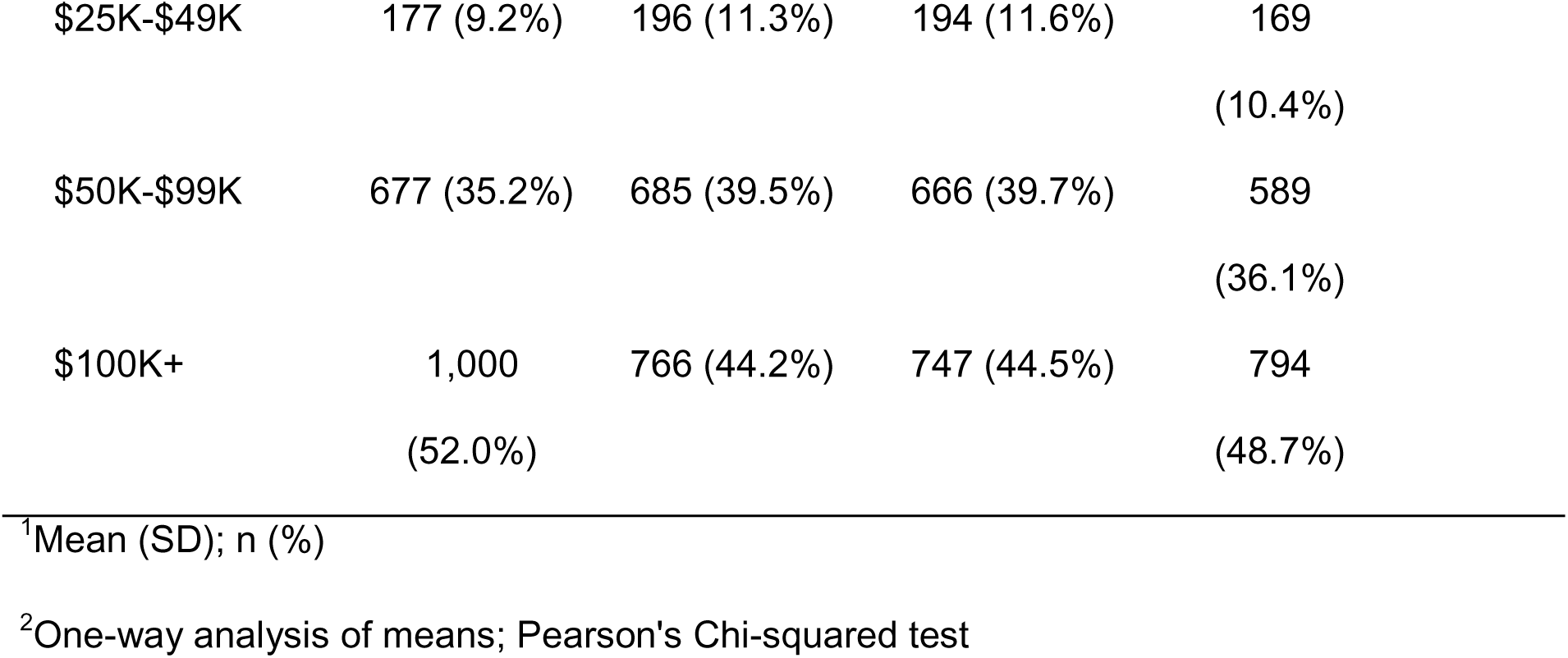
Sociodemographic Characteristics by Trajectory Cluster.

| Characteristic | Trajectory Cluster |  |  |  | p-value<sup>2</sup> |
| --- | --- | --- | --- | --- | --- |
|  | High-Stable | Declining | Persistently | Volatile |  |
|  | N = 1,922<sup>1</sup> | N = 1,734<sup>1</sup> | Low<br>N = 1,677<sup>1</sup> | N = 1,631<sup>1</sup> |  |
| Age at Baseline<br>(Months) | 119.15 (7.43) | 119.43<br>(7.48) | 119.62 (7.45) | 118.83<br>(7.50) | 0.013 |
| Sex |  |  |  |  | <0.001 |
| Male | 1,066<br>(55.5%) | 713 (41.1%) | 955 (56.9%) | 904<br>(55.4%) |  |
| Female | 856 (44.5%) | 1,021<br>(58.9%) | 722 (43.1%) | 727<br>(44.6%) |  |
| Race/Ethnicity |  |  |  |  | 0.009 |
| White | 1,212<br>(63.1%) | 997 (57.5%) | 994 (59.3%) | 997<br>(61.1%) |  |
| Hispanic | 320 (16.6%) | 347 (20.0%) | 325 (19.4%) | 287<br>(17.6%) |  |
| Black | 165 (8.6%) | 187 (10.8%) | 138 (8.2%) | 165<br>(10.1%) |  |
| Other/Mixed | 186 (9.7%) | 174 (10.0%) | 175 (10.4%) | 153 (9.4%) |  |
| Asian | 39 (2.0%) | 29 (1.7%) | 45 (2.7%) | 29 (1.8%) |  |
| Household<br>Income |  |  |  |  | <0.001 |
| < \$25K | 68 (3.5%) | 87 (5.0%) | 70 (4.2%) | 79 (4.8%) | |

|  |  |  |  |  |
| --- | --- | --- | --- | --- |
| \$25K-\$49K | 177 (9.2%) | 196 (11.3%) | 194 (11.6%) | 169<br>(10.4%) |
| \$50K-\$99K | 677 (35.2%) | 685 (39.5%) | 666 (39.7%) | 589<br>(36.1%) |
| \$100K+ | 1,000<br>(52.0%) | 766 (44.2%) | 747 (44.5%) | 794<br>(48.7%) |

<sup>1</sup>Mean (SD); n (%)
<sup>2</sup>One-way analysis of means; Pearson's Chi-squared test

### Multinomial regression

Models revealed significant associations between the ELA composite score and positive affect trajectory membership. With the High and Stable group serving as the reference category, higher ELA scores were significantly associated with membership in all other trajectory groups. The strongest association was observed for the Persistently Low trajectory group (β = 0.321, z = 7.99, p < 0.001), followed by the Volatile trajectory group (β = 0.171, z = 4.00, p < 0.001) and the Declining trajectory group (β = 0.144, z = 3.34, p < 0.001) in models controlling for race/ethnicity, initial interview age (in months), sex assigned at birth, and recruitment site. Figure 2 displays the predicted probabilities of trajectory group membership across the range of ELA scores, illustrating that as ELA exposure increases, the probability of belonging to the High and Stable group decreases while probabilities of membership in the Persistently Low, Volatile, and Declining groups increase. Among youth with minimal adversity exposure, group membership was relatively evenly distributed. However, as adversity increased, the Declining trajectory emerged as the dominant pathway, increasing from 20% probability at low adversity to approximately 65-70% at the highest adversity levels. Conversely, probability of High and Stable membership dropped to 4-8.5% at high adversity. Results were similar for sensitivity models controlling for socioeconomic status, with ELA still relating to membership in the Declining trajectory group (β = 0.101, z = 2.22, p = 0.026), Persistently Low trajectory (β = 0.300, z = 7.04, p < 0.001), and Volatile trajectory (β = 0.146, z = 3.23, p = 0.001).

**Figure 2.**
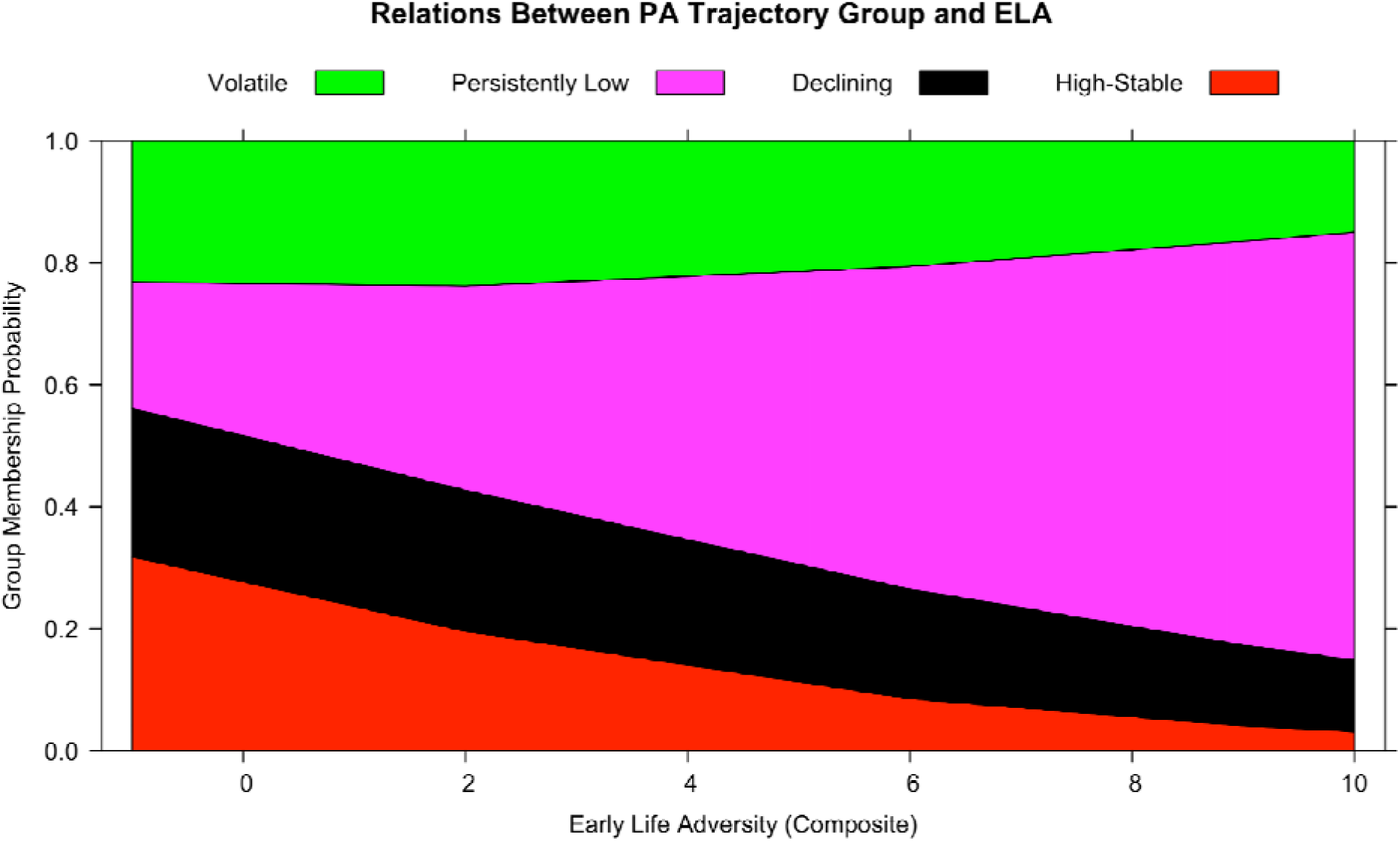
**Caption:** Relationship between positive affect trajectory group membership probability and early life adversity (ELA) composite scores. The stacked area chart displays the predicted probabilit of membership in four trajectory groups: Volatile (green), Persistently Low (magenta), Declining (black), and High-Stable (red). As early life adversity increases, the probability of High-Stable positive affect decreases while the probability of Persistently Low and Volatile patterns increases.

### Mediation

We lastly examined if positive affect trajectory membership mediated links between ELA and internalizing or externalizing psychopathology. While ELA was directly related to externalizing issues (β = 0.338, z = 3.11, p = 0. 002), none of the individual trajectory pathways showed significant mediation effects for externalizing problems. The indirect effects through the Declining, Persistently Low, and Volatile trajectory groups were all non-significant (all p’s>0.101). For internalizing problems, ELA was directly related to internalizing issues (β = 0.248, z = 2.23, p = 0.026) and significant (partial) mediation was observed through the Persistently Low trajectory group (indirect effect = 0.030, 95% CI [0.012, 0.056], z = 2.679, p = 0.007). However, the indirect effects through the Declining and Volatile trajectory groups were not significant (p’s>0.485). **Figure 3** presents the indirect effects of ELA on externalizing and internalizing problems through each positive affect trajectory group.^1^ **Table 3** lists the standardized coefficients with standard errors for relations between ELA and trajectory group, trajectory group and psychopathology, and ELA and psychopathology.^2^ Similar effects were found in sensitivity models controlling socioeconomic status. No cluster membership mediated relations between ELA and externalizing psychopathology symptoms (all p’s>0.106). For internalizing, when controlling for socioeconomic status, significant mediation was observed through the Persistently Low trajectory group (indirect effect = 0.033, 95% CI [0.013, 0.058], z = 2.747, p = 0.006).

**Figure 3.**
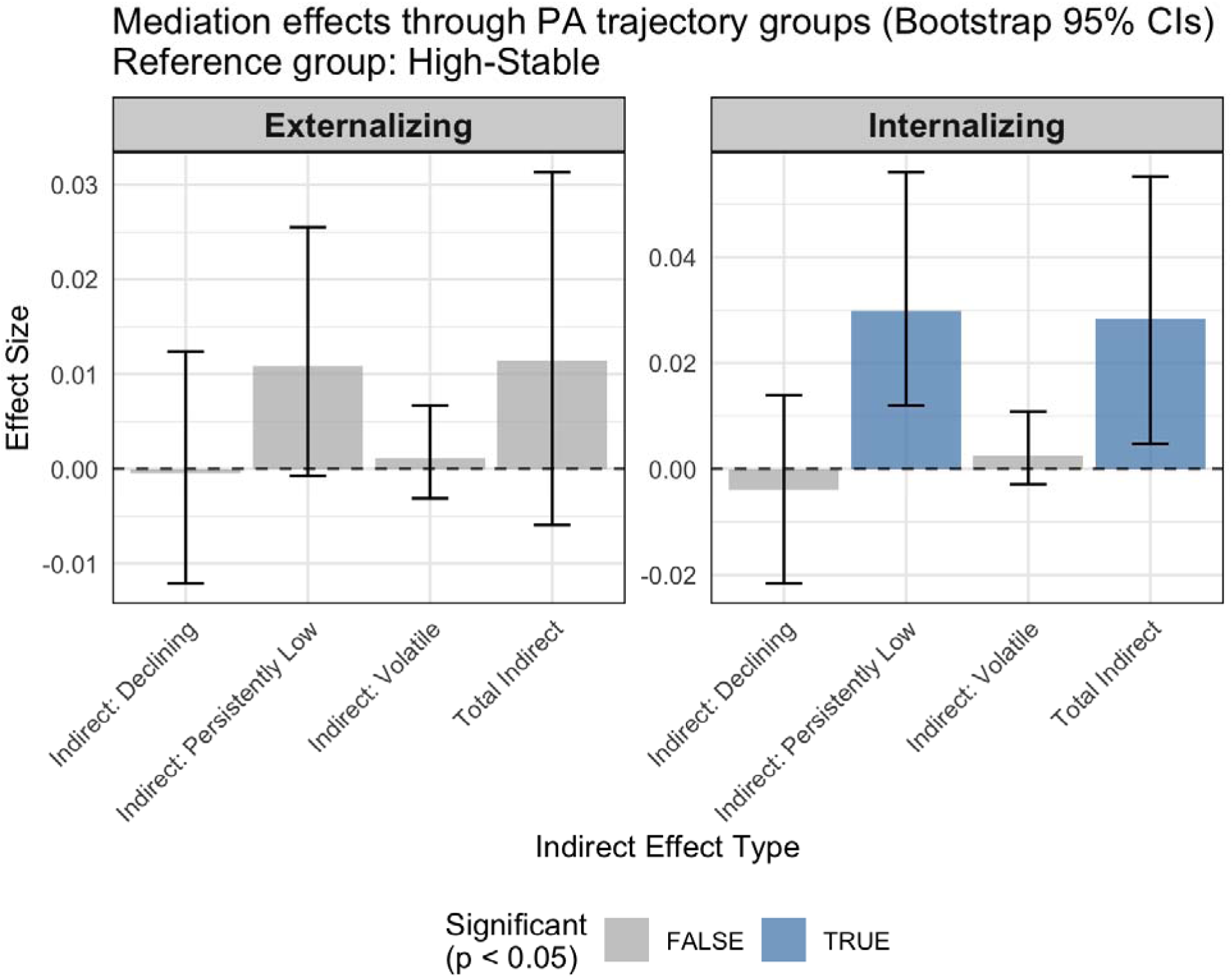
**Caption**: Mediation effects of positive affect trajectory groups on externalizing and internalizing outcomes with bootstrap 95% confidence intervals. The High-Stable trajectory group serves as the reference category. Standardized effect sizes are shown for the indirect effect (ELA -> Group X Group -> Psychopathology symptoms) for each group (declining, persistently low, volatile). The total indirect effects are shown on the right portion of each panel. Blue bars indicate statistically significant effects (p < 0.05), while gray bars represent non-significant effects. Significant mediation effects are observed for internalizing outcomes through persistently low positive affect trajectory pathways.

**Table 3.** Mediation Analysis: Path Coefficients^1^ Standardized coefficients with standard errors^1^. Standardized coefficients with standard errors^1^

| Path | Internalizing<br>Problems<sup>2</sup> |  | Externalizing<br>Problems<sup>2</sup> |  |
| --- | --- | --- | --- | --- |
| | $\beta$ | SE | $\beta$ | SE |
| **a paths** |  |  |  |  |
| ELA → Declining | -0.007 | (0.008) | -0.001 | (0.008) |
| ELA → Persistently Low | 0.062*** | (0.008) | 0.062*** | (0.008) |
| ELA → Volatile | 0.021 | (0.009) | 0.019 | (0.009) |
| **b paths** |  |  |  |  |
| Declining → Outcome | 0.079*** | (0.222) | 0.061*** | (0.190) |
| Persistently Low →<br>Outcome | 0.070*** | (0.223) | 0.030 | (0.189) |
| Volatile → Outcome | 0.017 | (0.207) | 0.010 | (0.180) |
| **c' path** |  |  |  |  |
| ELA → Outcome (Direct) | 0.036* | (0.111) | 0.058** | (0.109) |

<sup>1</sup>Reference group: High-Stable positive affect trajectory<sup>2</sup>\*\*\* p < 0.001, \*\* p < 0.01, \* p < 0.05

## Discussion

In a large, nationally representative sample, we found that ELA was related to different trajectories of positive affect across childhood and adolescence, which in turn predicted internalizing psychopathology. Adversity experienced in childhood was related to a child’s likelihood of having a low, declining, or volatile trajectory of positive affect compared to high and stable positive affect trajectory. In fact, as adversity increases, the probability of being in the high and stable positive affect group was very low (4-8.5%). Notably, the persistently low positive affect trajectory group was strongly associated with ELA and statistically explained the link between ELA and later internalizing symptoms. The person-centered approach used here allowed us to isolate unique developmental trajectories that may mark risk or resilience after ELA, moving away from variable-centered models (i.e., examining mean group differences) that have dominated past explorations.

Results revealed that positive emotions, or a lack thereof, may be an important mechanism linking ELA to later internalizing symptomatology including anxiety, depression, and withdrawal. Previous work on affect-related impacts of ELA have uncovered alterations in threat and negative affect processing (Gorka et al., 2014; Hanson, Nacewicz, et al., 2015; Hanson & Nacewicz, 2021; Kraynak et al., 2019), but such a focus on negative emotions discounts the fact that reduced positive emotions in the context of stress may also increase risk for depression and anxiety (Khazanov & Ruscio, 2016; Sequeira et al., 2022). The current findings align with emerging theories suggesting that ELAs disrupt core emotional and regulatory systems during sensitive developmental windows, including attachment processes, neuroendocrine functioning, and the maturation of brain regions involved in learning, memory, and behavioral control (Kennedy et al., 2021; Palacios-Barrios et al., 2024; Shirtcliff et al., 2024). Notably, we found that positive affect statistically mediated links between ELA and internalizing – but not externalizing – problems, consistent with previous research finding that those experiencing low positive affect after stress are much more likely to develop later depression (Rackoff & Newman, 2020). Such findings underscore the possibility that ELA-related disruptions in the development of positive affect may *specifically* heighten vulnerability to internalizing problems such as depression and anxiety, rather than contributing *broadly* to multiple forms of psychopathology. This differential pathway specificity has important implications for understanding heterogeneity in mental health outcomes following ELA.

The heterogeneity in empirically derived positive affect trajectories may reflect differences in the emotion regulatory and neural circuits supporting positive emotional experiences. Meta-analyses of group-level findings (i.e., statistics examining mean differences) have found that ELA is related to lower positive affect (Lavi et al., 2019), impaired reward processing (Oltean et al., 2023), and lower responsivity in neural circuits critical for reward processing (Birn et al., 2017; Hanson et al., 2016; Hanson, Hariri, et al., 2015; Sacu et al., 2024; van Harmelen et al., 2014); for review, see (Hanson et al., 2021). However, group-level statistical approaches may mask important subtypes of trajectories after adversity. Future work could delve into neurobiological heterogeneity to differentiate subtypes of brain reactivity and functional connectivity in those exposed to ELA and examine whether and how such subtypes predict depression and anxiety symptomatology.

This study benefited from numerous strengths and points to several promising directions for future research. The large, geographically diverse sample enhances the generalizability of findings across populations and contexts, while the longitudinal design – spanning a sensitive developmental period from childhood through adolescence – captures a window of rapid emotional and neurobiological change. Although many youth exposed to ELA showed persistently low or declining positive affect, others maintained high levels of positive affect despite having experienced significant adversity. One critical next step for research is identifying the factors that support these more adaptive emotional trajectories. Recent work has highlighted certain neurobiological features such as variations in brain structure and functional connectivity as potential protective factors that predict positive adaptation across life domains (Hanson et al., 2019; Suarez et al., 2025). Future research should explore whether these neurobiological and psychosocial characteristics function as resilience markers, helping to buffer the impact of ELA and support mental health across development.

While our work has several strengths, there are several limitations to highlight. First, our approach to modeling ELA involved several methodological decisions that warrant discussion. We employed a continuous z-score composite that averaged items within measurement scales, z-scored at the scale level, and then averaged across scales. This approach offers several advantages, including greater sensitivity to variation in adversity levels and a parsimonious single measure that captures cumulative risk burden. However, like all cumulative risk approaches, our measure assumes equal impact of each adverse event and does not account for factors such as timing, severity, or individual perception of stress (Smith & Pollak, 2021). Alternative approaches, such as binary domain scoring where each adversity domain (e.g., physical abuse, emotional neglect) is coded as present or absent regardless of the number of items measuring that domain, may reduce measurement bias when domains are assessed with differing numbers of items (Breslin et al., 2025). Such approaches also allow for creation of multiple versions tailored to specific research questions, such as measures focused on traditional ACEs domains versus broader ELA conceptualizations that include exposure to community violence, lack of resources, and caregiver separation. While our continuous composite approach was well-suited to examining dose-response relationships between adversity and developmental trajectories, future research could benefit from comparing findings across different operationalizations of ELA to understand how measurement choices influence conclusions about risk and resilience. Moreover, future research could incorporate dimensional ELA models and center youth perspectives on their subjective experiences to better understand how different ELA facets affect mental health (Kahhalé et al., 2023; A. B. Miller et al., 2018; P. Miller et al., 2025). Second, while the study assessments span several years, extended follow-up periods into adulthood would provide important information about the persistence of observed patterns and their longer-term mental health consequences. Third, the positive affect measure asked participants to reflect on feelings over the past week, which captures a shorter timeframe than measures assessing affect “in general”. Additionally, adolescent affect is known to fluctuate considerably in response to various situational factors (Reitsema et al., 2022). While this could raise questions about stability versus transient states, past evidence supports the stability and meaningfulness of these measurements (Diener et al., 2018; Watson, 1988) and our ability to consistently classify trajectory groups suggests these patterns capture meaningful developmental differences. Nonetheless, future research could examine convergence with daily diary methods, experience sampling approaches, or trait-level affect measures to further characterize sources of variance and better distinguish developmental shifts from situational influences. Fourth, while positive affect was the focus of the current study, future studies should examine how ELA affects the balance between positive and negative emotional systems. The volatile trajectory group, in particular, may reflect disruptions in emotion regulation that affect both positive and negative emotional responses. Finally, as noted in Appendix *S4-S5*, data was not missing at random. However, we believe this missingness could actually be underestimating our effects of interest, as excluded participants may be drawn from various “high risk” demographics (e.g., lower income; families of color who face structural adversity, higher ELA).

Our work provides novel evidence that ELA shapes positive affect development in heterogeneous ways, with important implications for understanding risk and resilience pathways to depression and anxiety. The finding that persistently low positive affect partially mediates the connection between ELA and internalizing symptoms suggests a viable target for interventions. Early identification of persistently low positive affect may enable treatments focused on enhancing positive emotional experiences and building resilience before depressive symptoms fully emerge (Taylor et al., 2017). This work and future studies deploying nuanced, theory-driven modeling of heterogeneity across ELA, affect, and psychopathology could identify vulnerable youth and optimize timing and targets for intervention.

## Supporting information

Supplemental Materials/Analyses

## Abbreviations

ELAs: Early life adversities
ABCD: Adolescent Brain Cognitive Development Study

## Acknowledgments

The ABCD consortium did not participate in the analysis or writing of this report. This article reflects the views of the authors and may not reflect the opinions or views of NIH or of the ABCD consortium investigators. This work was supported by the National Institute of Mental Health under grant R01MH136109 to J.H., as well as internal funding from the University of Pittsburgh and Pitt’s Learning Research & Development Center provided to J.H. The authors have declared that they have no competing or potential conflicts of interest.

## Data Availability Statement

Data used in this study were obtained from the Adolescent Brain Cognitive Development (ABCD) Study (https://abcdstudy.org), held in the NIMH Data Archive. The ABCD data used in this report came from the 5.1 release (information available here: https://doi.org/10.15154/cqdy-5453). Relevant preprocessing scripts will be made available via a Github repository upon publication of the work.

## Key Points

### What’s known?

Early life adversity (ELA) is established as a risk factor for depression, but prior research has predominantly focused on negative emotions rather than examining how ELA shapes positive emotional development across childhood and adolescence.

### What’s new?

Using person-centered trajectory modeling in 7,457 youth, we identified four distinct positive affect patterns: High-Stable (27.6%), Declining (24.8%), Persistently Low (24.4%), and Volatile (23.3%). ELA exposure significantly predicted membership in all non-stable trajectory groups. Critically, persistently low positive affect specifically mediated the relationship between ELA and internalizing problems (depression/anxiety) but not externalizing problems, revealing a targeted pathway to specific forms of psychopathology.

### What’s relevant?

Early identification of youth with persistently low positive affect trajectories could enable targeted interventions focused on enhancing positive emotional experiences before depressive symptoms fully emerge.

## Supporting Information

Appendix S1. Measurement of Early Life Adversity

Appendix S2. Trajectory Features

Appendix S3. Sensitivity Analysis: Mediation Using Alternative Formulations of Internalizing Subscales

Appendix S4. Missing Data Analysis: Trajectory Analyses

Appendix S5. Missing Data Analysis: Mediation Analyses

Table S1. Descriptions for Each ELA Item

Table S2: Trajectory Features Used in Clustering Analysis

Table S3: Items Removed: Reduced Overlap 21-Item Scale

Figure S1. Distribution of ELA Z-score

Figure S2. Mediation Using Alternative Formulations of Internalizing Subscales

Figure S3. Missing Data Diagram

## Footnotes

1 The High and Stable group again served as the reference category.

2 Analyses were completed in 3,927 participants, as 3,530 participants did not have data from the CBCL at the 4-year follow-up timepoint of the ABCD study.

## References

Achenbach, T. M. (2011). Child behavior checklist. In Encyclopedia of clinical neuropsychology (pp. 546–552). Springer.

Alexander, R., Aragón, O. R., Bookwala, J., Cherbuin, N., Gatt, J. M., Kahrilas, I. J., Kästner, N., Lawrence, A., Lowe, L., Morrison, R. G., & others. (2021). The neuroscience of positive emotions and affect: Implications for cultivating happiness and wellbeing. Neuroscience & Biobehavioral Reviews, 121, 220–249.

Bailen, N. H., Green, L. M., & Thompson, R. J. (2019). Understanding emotion in adolescents: A review of emotional frequency, intensity, instability, and clarity. Emotion Review, 11(1), 63–73.

Bhutta, Z. A., Bhavnani, S., Betancourt, T. S., Tomlinson, M., & Patel, V. (2023). Adverse childhood experiences and lifelong health. Nature Medicine, 29(7), 1639–1648.

Bick, J., Luyster, R., Fox, N. A., Zeanah, C. H., & Nelson, C. A. (2017). Effects of early institutionalization on emotion processing in 12-year-old youth. Development and Psychopathology, 29(5), 1749–1761.

Birn, R. M., Roeber, B. J., & Pollak, S. D. (2017). Early childhood stress exposure, reward pathways, and adult decision making. Proceedings of the National Academy of Sciences, 114(51), 13549–13554.

Bunea, I. M., Szentágotai-Tătar, A., & Miu, A. C. (2017). Early-life adversity and cortisol response to social stress: A meta-analysis. Translational Psychiatry, 7(12), 1274.

Bylsma, L. M., Morris, B. H., & Rottenberg, J. (2008). A meta-analysis of emotional reactivity in major depressive disorder. Clinical Psychology Review, 28(4), 676–691.

Caliński, T., & Harabasz, J. (1974). A dendrite method for cluster analysis. Communications in Statistics-Theory and Methods, 3(1), 1–27.

Caouette, J., Cossette, L., & Hébert, M. (2023). Do you see what I see? Emotion recognition competencies in sexually abused school-aged children and non-abused children. Journal of Child Sexual Abuse, 32(7), 813–828.

Clark, L. A., Watson, D., & Leeka, J. (1989). Diurnal variation in the positive affects. Motivation and Emotion, 13, 205–234.

Colman, I., Kingsbury, M., Garad, Y., Zeng, Y., Naicker, K., Patten, S., Jones, P. B., Wild, T. C., & Thompson, A. H. (2016). Consistency in adult reporting of adverse childhood experiences. Psychological Medicine, 46(3), 543–549.

Cyr, C., Euser, E. M., Bakermans-Kranenburg, M. J., & Van Ijzendoorn, M. H. (2010). Attachment security and disorganization in maltreating and high-risk families: A series of meta-analyses. Development and Psychopathology, 22(1), 87–108.

Dadds, M. R., Mullins, M. J., McAllister, R. A., & Atkinson, E. (2003). Attributions, affect, and behavior in abuse-risk mothers: A laboratory study. Child Abuse & Neglect, 27(1), 21–45. 10.1016/S0145-2134(02)00510-0

DePierro, J., D’Andrea, W., Frewen, P., & Todman, M. (2018). Alterations in positive affect: Relationship to symptoms, traumatic experiences, and affect ratings. *Psychological Trauma: Theory, Research*, Practice, and Policy, 10(5), 585–593. 10.1037/tra0000317

Diaconu, B., Kohls, G., Rogers, J. C., Pauli, R., Cornwell, H., Bernhard, A., Martinelli, A., Ackermann, K., Fann, N., Fernandez-Rivas, A., & others. (2023). Emotion processing in maltreated boys and girls: Evidence for latent vulnerability. European Child & Adolescent Psychiatry, 32(12), 2523–2536.

Dillon, D. G., Holmes, A. J., Birk, J. L., Brooks, N., Lyons-Ruth, K., & Pizzagalli, D. A. (2009). Childhood adversity is associated with left basal ganglia dysfunction during reward anticipation in adulthood. Biological Psychiatry, 66(3), 206–213.

Doyle, C., & Cicchetti, D. (2017). From the cradle to the grave: The effect of adverse caregiving environments on attachment and relationships throughout the lifespan. Clinical Psychology: Science and Practice, 24(2), 203.

Egeland, B., & Sroufe, A. (1981). Developmental sequelae of maltreatment in infancy. New Directions for Child and Adolescent Development, 1981(11), 77–92. 10.1002/cd.23219811106

Egeland, B., Sroufe, L. A., & Erickson, M. (1983). The developmental consequence of different patterns of maltreatment. Child Abuse & Neglect, 7(4), 459–469. 10.1016/0145-2134(83)90053-4

Etter, D. W., Gauthier, J. R., McDade-Montez, E., Cloitre, M., & Carlson, E. B. (2013). Positive affect, childhood adversity, and psychopathology in psychiatric inpatients. European Journal of Psychotraumatology, 4(1), 20771. 10.3402/ejpt.v4i0.20771

Felitti, V. J., Anda, R. F., Nordenberg, D., Williamson, D. F., Spitz, A. M., Edwards, V., & Marks, J. S. (1998). Relationship of childhood abuse and household dysfunction to many of the leading causes of death in adults: The Adverse Childhood Experiences (ACE) Study. American Journal of Preventive Medicine, 14(4), 245–258.

Folkman, S. (2008). The case for positive emotions in the stress process. *Anxiety*, Stress, and Coping, 21(1), 3–14.

Fries, A. B. W., & Pollak, S. D. (2004). Emotion understanding in postinstitutionalized Eastern European children. Development and Psychopathology, 16(2), 355–369.

Gee, D. G. (2021). Early adversity and development: Parsing heterogeneity and identifying pathways of risk and resilience. American Journal of Psychiatry, 178(11), 998–1013.

Gilgoff, R., Singh, L., Koita, K., Gentile, B., & Marques, S. S. (2020). Adverse childhood experiences, outcomes, and interventions. Pediatric Clinics, 67(2), 259–273.

Glaser, J.-P., Van Os, J., Portegijs, P. J., & Myin-Germeys, I. (2006). Childhood trauma and emotional reactivity to daily life stress in adult frequent attenders of general practitioners. Journal of Psychosomatic Research, 61(2), 229–236.

Gorka, A. X., Hanson, J. L., Radtke, S. R., & Hariri, A. R. (2014). Reduced hippocampal and medial prefrontal gray matter mediate the association between reported childhood maltreatment and trait anxiety in adulthood and predict sensitivity to future life stress. Biology of Mood & Anxiety Disorders, 4, 1–10.

Green, J. G., McLaughlin, K. A., Berglund, P. A., Gruber, M. J., Sampson, N. A., Zaslavsky, A. M., & Kessler, R. C. (2010). Childhood adversities and adult psychiatric disorders in the national comorbidity survey replication I: associations with first onset of DSM-IV disorders. Archives of General Psychiatry, 67(2), 113–123.

Hair, N. L., Hanson, J. L., Wolfe, B. L., & Pollak, S. D. (2015). Association of child poverty, brain development, and academic achievement. JAMA Pediatrics, 169(9), 822–829.

Hanson, J. L., Albert, D., Iselin, A.-M. R., Carre, J. M., Dodge, K. A., & Hariri, A. R. (2016). Cumulative stress in childhood is associated with blunted reward-related brain activity in adulthood. Social Cognitive and Affective Neuroscience, 11(3), 405–412.

Hanson, J. L., Chandra, A., Wolfe, B. L., & Pollak, S. D. (2011). Association between income and the hippocampus. PloS One, 6(5), e18712.

Hanson, J. L., Chung, M. K., Avants, B. B., Rudolph, K. D., Shirtcliff, E. A., Gee, J. C., Davidson, R. J., & Pollak, S. D. (2012). Structural variations in prefrontal cortex mediate the relationship between early childhood stress and spatial working memory. Journal of Neuroscience, 32(23), 7917–7925.

Hanson, J. L., Chung, M. K., Avants, B. B., Shirtcliff, E. A., Gee, J. C., Davidson, R. J., & Pollak, S. D. (2010). Early stress is associated with alterations in the orbitofrontal cortex: A tensor-based morphometry investigation of brain structure and behavioral risk. Journal of Neuroscience, 30(22), 7466–7472.

Hanson, J. L., Gillmore, A. D., Yu, T., Holmes, C. J., Hallowell, E. S., Barton, A. W., Beach, S. R., Galván, A., MacKillop, J., Windle, M., & others. (2019). A family focused intervention influences hippocampal-prefrontal connectivity through gains in self-regulation. Child Development, 90(4), 1389–1401.

Hanson, J. L., Hair, N., Shen, D. G., Shi, F., Gilmore, J. H., Wolfe, B. L., & Pollak, S. D. (2013). Family poverty affects the rate of human infant brain growth. PloS One, 8(12), e80954.

Hanson, J. L., Hariri, A. R., & Williamson, D. E. (2015). Blunted ventral striatum development in adolescence reflects emotional neglect and predicts depressive symptoms. Biological Psychiatry, 78(9), 598–605.

Hanson, J. L., Kahhalé, I., & Sen, S. (2024). Integrating data science and neuroscience in developmental psychopathology: Formative examples and future directions. Development and Psychopathology, 36(5), 2165–2172.

Hanson, J. L., & Nacewicz, B. M. (2021). Amygdala allostasis and early life adversity: Considering excitotoxicity and inescapability in the sequelae of stress. Frontiers in Human Neuroscience, 15, 624705.

Hanson, J. L., Nacewicz, B. M., Sutterer, M. J., Cayo, A. A., Schaefer, S. M., Rudolph, K. D., Shirtcliff, E. A., Pollak, S. D., & Davidson, R. J. (2015). Behavioral problems after early life stress: Contributions of the hippocampus and amygdala. Biological Psychiatry, 77(4), 314–323.

Hanson, J. L., van den Bos, W., Roeber, B. J., Rudolph, K. D., Davidson, R. J., & Pollak, S. D. (2017). Early adversity and learning: Implications for typical and atypical behavioral development. Journal of Child Psychology and Psychiatry, 58(7), 770–778.

Hanson, J. L., Williams, A. V., Bangasser, D. A., & Peña, C. J. (2021). Impact of early life stress on reward circuit function and regulation. Frontiers in Psychiatry, 12, 744690.

Hill, P. L., Turiano, N. A., & Burrow, A. L. (2018). Early life adversity as a predictor of sense of purpose during adulthood. International Journal of Behavioral Development, 42(1), 143–147.

Humphreys, K. L., LeMoult, J., Wear, J. G., Piersiak, H. A., Lee, A., & Gotlib, I. H. (2020). Child maltreatment and depression: A meta-analysis of studies using the Childhood Trauma Questionnaire. Child Abuse & Neglect, 102, 104361.

Kahhalé, I., Barry, K. R., & Hanson, J. L. (2023). Positive parenting moderates associations between childhood stress and corticolimbic structure. PNAS Nexus, 2(6), pgad145.

Karcher, N. R., & Barch, D. M. (2021). The ABCD study: Understanding the development of risk for mental and physical health outcomes. Neuropsychopharmacology, 46(1), 131–142.

Kennedy, B. V., Hanson, J. L., Buser, N. J., van den Bos, W., Rudolph, K. D., Davidson, R. J., & Pollak, S. D. (2021). Accumbofrontal tract integrity is related to early life adversity and feedback learning. Neuropsychopharmacology, 46(13), 2288–2294.

Khazanov, G. K., & Ruscio, A. M. (2016). Is low positive emotionality a specific risk factor for depression? A meta-analysis of longitudinal studies. Psychological Bulletin, 142(9), 991.

Koizumi, M., & Takagishi, H. (2014). The relationship between child maltreatment and emotion recognition. PloS One, 9(1), e86093.

Kraynak, T. E., Marsland, A. L., Hanson, J. L., & Gianaros, P. J. (2019). Retrospectively reported childhood physical abuse, systemic inflammation, and resting corticolimbic connectivity in midlife adults. *Brain*, Behavior, and Immunity, 82, 203–213.

Kuzminskaite, E., Vinkers, C. H., Smit, A. C., Ballegooijen, W. van, Elzinga, B. M., Riese, H., Milaneschi, Y., & Penninx, B. W. J. H. (2024). Day-to-day affect fluctuations in adults with childhood trauma history: A two-week ecological momentary assessment study. Psychological Medicine, 54(6), 1160–1171. 10.1017/S0033291723002969

Larson, R. W., Moneta, G., Richards, M. H., & Wilson, S. (2002). Continuity, stability, and change in daily emotional experience across adolescence. Child Development, 73(4), 1151–1165.

Lavi, I., Katz, L. F., Ozer, E. J., & Gross, J. J. (2019). Emotion reactivity and regulation in maltreated children: A meta-analysis. Child Development, 90(5), 1503–1524.

Leffondré, K., Abrahamowicz, M., Regeasse, A., Hawker, G. A., Badley, E. M., McCusker, J., & Belzile, E. (2004). Statistical measures were proposed for identifying longitudinal patterns of change in quantitative health indicators. Journal of Clinical Epidemiology, 57(10), 1049–1062.

LeMoult, J., Humphreys, K. L., Tracy, A., Hoffmeister, J.-A., Ip, E., & Gotlib, I. H. (2020). Meta-analysis: Exposure to early life stress and risk for depression in childhood and adolescence. Journal of the American Academy of Child & Adolescent Psychiatry, 59(7), 842–855.

Linden, A. H., & Hönekopp, J. (2021). Heterogeneity of research results: A new perspective from which to assess and promote progress in psychological science. Perspectives on Psychological Science, 16(2), 358–376.

Madigan, S., Deneault, A.-A., Racine, N., Park, J., Thiemann, R., Zhu, J., Dimitropoulos, G., Williamson, T., Fearon, P., Cénat, J. M., & others. (2023). Adverse childhood experiences: A meta-analysis of prevalence and moderators among half a million adults in 206 studies. World Psychiatry, 22(3), 463–471.

Mandelli, L., Petrelli, C., & Serretti, A. (2015). The role of specific early trauma in adult depression: A meta-analysis of published literature. Childhood trauma and adult depression. European Psychiatry, 30(6), 665–680.

McLaughlin, K. A., Hatzenbuehler, M. L., & Hilt, L. M. (2009). Emotion dysregulation as a mechanism linking peer victimization to internalizing symptoms in adolescents. Journal of Consulting and Clinical Psychology, 77(5), 894.

Melamed, D. M., Botting, J., Lofthouse, K., Pass, L., & Meiser-Stedman, R. (2024). The relationship between negative self-concept, trauma, and maltreatment in children and adolescents: A meta-analysis. Clinical Child and Family Psychology Review, 1–15.

Miller, A. B., Sheridan, M. A., Hanson, J. L., McLaughlin, K. A., Bates, J. E., Lansford, J. E., Pettit, G. S., & Dodge, K. A. (2018). Dimensions of deprivation and threat, psychopathology, and potential mediators: A multi-year longitudinal analysis. Journal of Abnormal Psychology, 127(2), 160.

Miller, P., Blatt, L., Hunter-Rue, D., Barry, K. R., Jamal-Orozco, N., Hanson, J. L., & Votruba-Drzal, E. (2025). Economic hardship and adolescent behavioral outcomes: Within-and between-family associations. Development and Psychopathology, 37(1), 107–124.

Myroniuk, S., Reitsema, A. M., Jonge, P. de, & Jeronimus, B. F. (2024). Childhood abuse and neglect and profiles of adult emotion dynamics. Development and Psychopathology, 1–19. 10.1017/S0954579423001530

Nanni, V., Uher, R., & Danese, A. (2012). Childhood maltreatment predicts unfavorable course of illness and treatment outcome in depression: A meta-analysis. American Journal of Psychiatry, 169(2), 141–151.

Nweze, T., Ezenwa, M., Ajaelu, C., Hanson, J. L., & Okoye, C. (2023). Cognitive variations following exposure to childhood adversity: Evidence from a pre-registered, longitudinal study. Eclinicalmedicine, 56.

Oltean, L.-E., Loflău, R., Miu, A. C., & Szentágotai-Tătar, A. (2023). Childhood adversity and impaired reward processing: A meta-analysis. Child Abuse & Neglect, 142, 105596.

Palacios-Barrios, E. E., Patel, K., & Hanson, J. L. (2024). Early life interpersonal stress and depression: Social reward processing as a potential mediator. Progress in Neuro-Psychopharmacology and Biological Psychiatry, 129, 110887.

Pears, K. C., & Fisher, P. A. (2005). Emotion understanding and theory of mind among maltreated children in foster care: Evidence of deficits. Development and Psychopathology, 17(1), 47–65.

Perlman, S. B., Kalish, C. W., & Pollak, S. D. (2008). The role of maltreatment experience in children’s understanding of the antecedents of emotion. Cognition & Emotion, 22(4), 651–670.

Pollak, S. D., & Kistler, D. J. (2002). Early experience is associated with the development of categorical representations for facial expressions of emotion. Proceedings of the National Academy of Sciences, 99(13), 9072–9076.

Rackoff, G. N., & Newman, M. G. (2020). Reduced positive affect on days with stress exposure predicts depression, anxiety disorders, and low trait positive affect 7 years later. Journal of Abnormal Psychology, 129(8), 799.

Reitsema, A. M., Jeronimus, B. F., van Dijk, M., & de Jonge, P. (2022). Emotion dynamics in children and adolescents: A meta-analytic and descriptive review. Emotion, 22(2), 374.

Robinson, L. R., Morris, A. S., Heller, S. S., Scheeringa, M. S., Boris, N. W., & Smyke, A. T. (2009). Relations Between Emotion Regulation, Parenting, and Psychopathology in Young Maltreated Children in Out of Home Care. Journal of Child and Family Studies, 18(4), 421–434. 10.1007/s10826-008-9246-6

Sacks, V., & Murphey, D. (2018). The prevalence of adverse childhood experiences, nationally, by state, and by race or ethnicity. Child Trends, 3.

Sacu, S., Dubois, M., Hezemans, F. H., Aggensteiner, P.-M., Monninger, M., Brandeis, D., Banaschewski, T., Hauser, T. U., & Holz, N. E. (2024). Early-life adversities are associated with lower expected value signaling in the adult brain. Biological Psychiatry, 96(12), 948–958.

Salsman, J. M., Butt, Z., Pilkonis, P. A., Cyranowski, J. M., Zill, N., Hendrie, H. C., Kupst, M. J., Kelly, M. A. R., Bode, R. K., Choi, S. W., Lai, J.-S., Griffith, J. W., Stoney, C. M., Brouwers, P., Knox, S. S., & Cella, D. (2013). Emotion assessment using the NIH Toolbox. Neurology, 80(11_supplement_3), S76–S86. 10.1212/WNL.0b013e3182872e11

Sequeira, S. L., Forbes, E. E., Hanson, J. L., & Silk, J. S. (2022). Positive valence systems in youth anxiety development: A scoping review. Journal of Anxiety Disorders, 89, 102588.

Shirtcliff, E. A., Hanson, J. L., Ruttle, P. L., Smith, B., & Pollak, S. D. (2024). Cortisol’s Diurnal Rhythm Indexes the Neurobiological Impact of Child Adversity in Adolescence. Biological Psychology, 108766.

Smith, K. E., & Pollak, S. D. (2021). Rethinking concepts and categories for understanding the neurodevelopmental effects of childhood adversity. Perspectives on Psychological Science, 16(1), 67–93.

Southwick, S. M., Vythilingam, M., & Charney, D. S. (2005). The psychobiology of depression and resilience to stress: Implications for prevention and treatment. Annu. Rev. Clin. Psychol., 1, 255–291.

Suarez, G. L., Bezek, J. L., Westerman, H. B., Hanson, J. L., Klump, K. L., Burt, S. A., & Hyde, L. W. (2025). Structural Brain Correlates of Multi-Domain Resilience Among Youth Exposed to Neighborhood Disadvantage. Biological Psychiatry Global Open Science, 100550.

Sylvestre, M.-P., & Vatnik, D. (2014). Using traj package to identify clusters of longitudinal trajectories. CRAN Packag. Traj, 593, 15.

Sylvestre, M.-P., Vatnik, D., & Vatnik, M. D. (2025). Package ‘traj.’

Taylor, C. T., Lyubomirsky, S., & Stein, M. B. (2017). Upregulating the positive affect system in anxiety and depression: Outcomes of a positive activity intervention. Depression and Anxiety, 34(3), 267–280.

Trøstheim, M., Eikemo, M., Meir, R., Hansen, I., Paul, E., Kroll, S. L., Garland, E. L., & Leknes, S. (2020). Assessment of anhedonia in adults with and without mental illness: A systematic review and meta-analysis. JAMA Network Open, 3(8), e2013233–e2013233.

Turiano, N. A., Silva, N. M., McDonald, C., & Hill, P. L. (2017). Retrospective reports of childhood misfortune are associated with positive and negative affect in adulthood: Exploring the moderating role of control beliefs. The International Journal of Aging and Human Development, 84(3), 276–293.

van Harmelen, A.-L., van Tol, M.-J., Dalgleish, T., van der Wee, N. J., Veltman, D. J., Aleman, A., Spinhoven, P., Penninx, B. W., & Elzinga, B. M. (2014). Hypoactive medial prefrontal cortex functioning in adults reporting childhood emotional maltreatment. Social Cognitive and Affective Neuroscience, 9(12), 2026–2033.

Volkow, N. D., Koob, G. F., Croyle, R. T., Bianchi, D. W., Gordon, J. A., Koroshetz, W. J., Pérez-Stable, E. J., Riley, W. T., Bloch, M. H., Conway, K., & others. (2018). The conception of the ABCD study: From substance use to a broad NIH collaboration. Developmental Cognitive Neuroscience, 32, 4–7.

Watson, D., Clark, L. A., & Tellegen, A. (1988). Development and validation of brief measures of positive and negative affect: The PANAS scales. Journal of Personality and Social Psychology, 54(6), 1063.

Wiersma, J. E., Hovens, J. G., van Oppen, P., Giltay, E. J., van Schaik, D. J., & Penninx, B. W. (2009). The importance of childhood trauma and childhood life events for chronicity of depression in adults. The Journal of Clinical Psychiatry, 70(7), 8733.

Williams, L. M., Debattista, C., Duchemin, A., Schatzberg, A. F., & Nemeroff, C. B. (2016). Childhood trauma predicts antidepressant response in adults with major depression: Data from the randomized international study to predict optimized treatment for depression. Translational Psychiatry, 6(5), e799–e799.

Wood, A. M., & Joseph, S. (2010). The absence of positive psychological (eudemonic) well-being as a risk factor for depression: A ten year cohort study. Journal of Affective Disorders, 122(3), 213–217.

Xiang, Y., Yuan, R., & Zhao, J. (2021). Childhood maltreatment and life satisfaction in adulthood: The mediating effect of emotional intelligence, positive affect and negative affect. Journal of Health Psychology, 26(13), 2460–2469. 10.1177/1359105320914381

Yarcheski, A., & Mahon, N. E. (2016). Meta-analyses of predictors of hope in adolescents. Western Journal of Nursing Research, 38(3), 345–368.

Yazgan, I., Hanson, J. L., Bates, J. E., Lansford, J. E., Pettit, G. S., & Dodge, K. A. (2021). Cumulative early childhood adversity and later antisocial behavior: The mediating role of passive avoidance. Development and Psychopathology, 33(1), 340–350.

Zhang, H., Wang, W., Liu, S., Feng, Y., & Wei, Q. (2023). A meta-analytic review of the impact of child maltreatment on self-esteem: 1981 to 2021. *Trauma, Violence*, & Abuse, 24(5), 3398–3411.

