## Supplemental Materials/Analyses for "Positive Affect as a Developmental Mediator of Early Adversity and Internalizing Psychopathology"

**Supporting Information**

***Appendix S1. Measurement of Early Life Adversity***

Measures of ELA were identified through examining ABCD documentation and literature (Brieant et al., 2023; Gong et al., 2021; Gonzalez et al., 2021; Hoffman et al., 2019; Orendain et al., 2023). Resulting items reflected categories of experiences captured in the original Adverse Childhood Experiences (ACEs) scale, a well-validated measure incorporated from the landmark ACEs study (Felitti et al., 1998). A standardized cumulative risk score was derived based on youth-reported and caregiver-reported items across a variety of scales administered at the baseline visit.

While multiple approaches exist for modeling ELA, cumulative risk scores have predictive power and allow researchers to consider exposure to multiple risk factors parsimoniously (Evans et al., 2013). Cumulative risk approaches also overcome many of the limitations to considering dimensions or specific types of adversities. These limitations to non-cumulative approaches include that boundaries differentiating adversity dimensions are ill-defined and often encompass multiple and overlapping types of experiences, that different adverse experiences tend to co-occur, and that subtypes of adversity lack specificity of effects on behavior and biology (Smith & Pollak, 2021). Taken together, these points suggest that a cumulative risk approach may be most appropriate to model the additive impact of ELA. Further, exposure to ELA in the present sample is assessed while participants are still in childhood, overcoming concerns (e.g., inaccurate memories) inherent to retrospective self-report on childhood adversity provided by participants when they are adults (Colman et al., 2016).

Our ELA composite score was derived from 42 items spanning eight ABCD Study questionnaires completed at baseline: the Kiddie Schedule for Affective Disorders and Schizophrenia (K-SADS), Family History Assessment, Neighborhood Safety and Crime, Parent Demographics Survey, Family Environment Scale, Youth Reported Parental Monitoring, Children's Report of Parent Behavior, and Parent Adult Self Report (see Table S1 for complete item list). Items captured multiple domains of adversity including: abuse and neglect (physical, sexual, emotional); exposure to community and domestic violence; household dysfunction (family conflict, poor monitoring, low caregiver warmth); material hardship (food insecurity, housing instability); neighborhood safety; and caregiver mental health concerns. Items were drawn from both youth self-report (n = 17 items) and parent report (n = 25 items) to provide comprehensive assessment across informants.

The methodology for creating the composite involved several steps. First, items within each scale were averaged for each participant. Several items (e.g., from the K-SADS) were already dichotomized (0 = no adversity, 1 = presence of adversity). For responses that were not binary (e.g., Likert scale responses), responses were z-scored at the scale-level. Next, scale averages were standardized across participants using z-scores. Finally, z-scores across all scales were averaged to generate a final averaged z-score representing cumulative risk for each participant. Any answer coded as "refuse to answer" or "don't know" was re-coded as missing data (NA). Only participants with at least 75% of ELA scales (minimum 6 of 8 scales) were included in the analysis. Higher scores reflect greater cumulative adversity exposure. Items are noted in Table S1 and a histogram showing this variable is shown in Figure S1. Among participants included in trajectory analyses (N = 7,262), the mean for this standardized composite was -0.09 (SD = 0.90) with a range from -1.09 to 12.61.

***Appendix S2. Trajectory Features***

The 19 trajectory features included measures of central tendency (e.g., mean, maximum), variability (e.g., standard deviation, range), linear trends (e.g., slope, intercept, R²), curvature (e.g., first and second derivatives), and temporal patterns (e.g., rate of intersection with mean, later/early change ratio). A complete description of all trajectory features is provided in Table S2.

***Appendix S3. Sensitivity Analysis: Mediation Using Alternative Formulations of Internalizing Subscales***

Sensitivity analyses using a 21-item internalizing composite (11 items removed shown in Table S3; r = .92 with original scale) yielded results consistent with the primary analysis. The total indirect effect through PA trajectories remained significant (β = 0.020, 95% CI [0.008, 0.035], p = .004), as did the specific pathway through Persistently Low PA trajectory (β = 0.020, 95% CI [0.009, 0.034], p = .003). Effect sizes were moderately smaller than in the original analysis (28-34% reductions), but the overall pattern of significance was unchanged. Results of this new analysis and the main analysis reported in the main manuscript are shown in Figure S3. These findings demonstrate that associations between PA trajectories and internalizing symptoms persist when items directly opposing positive affect are removed, supporting the interpretation that PA development predicts internalizing problems beyond shared measurement content.

***Appendix S4. Missing Data Analysis: Trajectory Analyses***

To assess whether data were missing completely at random (MCAR), we conducted Little's MCAR test, which indicated that data were not MCAR, χ²(28) = 207.00, p < .001. We compared participants included in trajectory analyses (N = 7,457) versus those excluded due to insufficient data (N = 4,411) on baseline characteristics. Participants excluded from analyses differed significantly from those included on several variables. Excluded participants had significantly lower baseline household income (M = 4.71, SD = 0.55) compared to included participants (M = 4.93, SD = 0.42), t(6441.1) = -21.95, p < .001. Race/ethnicity distributions also differed significantly, χ²(4) = 565.95, p < .001, with excluded participants more likely to identify as Black (24% vs. 10%) or Hispanic (22% vs. 19%), and less likely to identify as White (40% vs. 59%). Groups did not differ significantly on sex distribution (p = .50). Notably, excluded participants had significantly higher baseline ELA exposure (M = 0.15, SD = 1.13) compared to included participants (M = -0.09, SD = 0.90), t(7529.6) = 11.61, p < .001. Excluded participants were also slightly younger at baseline (M = 118.51 months, SD = 7.52) compared to included participants (M = 119.26 months, SD = 7.47), t(9206.3) = -5.28, p < .001.

***Appendix S5. Missing Data Analysis: Mediation Analyses***

To assess whether data were MCAR in the mediation sample, we conducted Little's MCAR test, which indicated that data were not MCAR, χ²(28) = 293.00, p < .001. We compared participants included in mediation analyses (N = 3,927) versus those excluded due to insufficient data (N = 7,941) on baseline characteristics. Participants excluded from analyses differed significantly from those included on several variables. Excluded participants had significantly lower baseline household income (M = 4.82, SD = 0.50) compared to included participants (M = 4.92, SD = 0.43), t(8557.4) = -11.37, p < .001. Race/ethnicity distributions also differed significantly, χ²(4) = 214.61, p < .001, with patterns similar to those observed in the trajectory sample. Groups did not differ significantly on sex distribution, χ²(2) = 0.32, p = .85. Notably, excluded participants had significantly higher baseline ELA exposure (M = 0.05, SD = 1.05) compared to included participants (M = -0.10, SD = 0.88), t(9276.7) = 7.96, p < .001. Excluded participants were also slightly younger at baseline (M = 118.71 months, SD = 7.51) compared to included participants (M = 119.52 months, SD = 7.44), t(7891.7) = -5.60, p < .001.

Of note, a flowchart showing sources of data loss is shown in Figure S3.

### **Table S1. Descriptions for Each ELA Item**

| Context | Construct | Survey | Item Description | Item Label | Informant |
| --- | --- | --- | --- | --- | --- |
| Neighborhood/ Environment | Natural Disaster | KSADS | Witnessed or caught in a natural disaster that caused significant property damage or personal injury? | ksads_ptsd_raw_757_p | Caregiver |
| Neighborhood/ Environment | Violence | KSADS | Witnessed or present during an act of terrorism (e.g., Boston marathon bombing)? | ksads_ptsd_raw_758_p | Caregiver |
| Neighborhood/ Environment | Violence | KSADS | Witnessed death or mass destruction in a war zone? | ksads_ptsd_raw_759_p | Caregiver |
| Neighborhood/  Environment | Violence | KSADS | Witnessed someone shot or stabbed in the community | ksads_ptsd_raw_760_p | Caregiver |
| Neighborhood/  Environment | Violence | KSADS | Shot, stabbed, or beaten brutally by a non-family member | ksads_ptsd_raw_761_p | Caregiver |
| Neighborhood/  Environment | Violence | KSADS | A non-family member threatened to kill your child | ksads_ptsd_raw_764_p | Caregiver |
| Neighborhood/  Environment | Violence | NSC | My neighborhood is safe from crime. | neighborhood3r_p | Caregiver |
| Neighborhood/  Environment | Violence | NSC | Violence is not a problem in my neighborhood. | neighborhood2r_p | Caregiver |
| Neighborhood/  Environment | Violence | NSC | My neighborhood is safe from crime. | neighborhood_crime_y | Youth |
| Neighborhood/  Environment | Sexual Abuse | KSADS | An adult outside your family touched your child in their privates, had your child touch their privates or did other sexual things to your child | ksads_ptsd_raw_768_p | Caregiver |
| Family/Home | Violence | KSADS | Shot, stabbed, or beaten brutally by a grown up in the home | ksads_ptsd_raw_762_p | Caregiver |
| Family/Home | Violence | KSADS | Beaten to the point of having bruises by a grown up in the home | ksads_ptsd_raw_763_p | Caregiver |
| Family/Home | Violence | KSADS | A family member threatened to kill your child | ksads_ptsd_raw_765_p | Caregiver |
| Family/Home | Violence | KSADS | Witness the grownups in the home push, shove or hit one another | ksads_ptsd_raw_766_p | Caregiver |
| Family/Home | Violence | FES | Family members sometimes hit each other. | fes_youth_q6 | Youth |
| Family/Home | Violence/ Emotional Abuse | FES | We fight a lot in our family. | fes_youth_q1 | Youth |
| Family/Home | Emotional Abuse | FES | Family members often criticize each other. | fes_youth_q5 | Youth |
| Family/Home | Sexual Abuse | KSADS | A grown up in the home touched your child in their privates, had your child touch their privates, or did other sexual things to your child? | ksads_ptsd_raw_767_p | Caregiver |
| Family/Home | Poverty | PDS | In the past 12 months, has there been a time when you and your immediate family experienced any of the following: Needed food but couldn't afford to buy it or couldn't afford to go out to get it? | demo_fam_exp1_v2 | Caregiver |
| Family/Home | Poverty | PDS | Had services turned off by the gas or electric company, or the oil company wouldn't deliver oil because payments were not made? | demo_fam_exp5_v2 | Caregiver |
| Family/Home | Poverty | PDS | In the past 12 months, has there been a time when you and your immediate family experienced any of the following: Were evicted from your home for not paying the rent or mortgage? | demo_fam_exp4_v2 | Caregiver |
| Family/Home | Caregiver Support/Monitoring | CRPB | First caregiver makes me feel better after talking over my worries with him/her | crpbi_parent1_y | Youth |
| Family/Home | Caregiver Support/Monitoring | CRPB | First caregiver smiles at me very often. | crpbi_parent2_y | Youth |
| Family/Home | Caregiver Support/Monitoring | CRPB | First caregiver is able to make me feel better when I am upset. | crpbi_parent3_y | Youth |
| Family/Home | Caregiver Support/Monitoring | CRPB | First caregiver believes in showing his/her love for me. | crpbi_parent4_y | Youth |
| Family/Home | Caregiver Support/Monitoring | CRPB | First caregiver is easy to talk to. | crpbi_parent5_y | Youth |
| Family/Home | Caregiver Support/Monitoring | CRPB | Second caregiver makes me feel better after talking over my worries with them. | crpbi_caregiver12_y | Youth |
| Family/Home | Caregiver Support/Monitoring | CRPB | Second caregiver smiles at me very often. | crpbi_caregiver13_y | Youth |
| Family/Home | Caregiver Support/Monitoring | CRPB | Second caregiver is able to make me feel better when I am upset. | crpbi_caregiver14_y | Youth |
| Family/Home | Caregiver Support/Monitoring | CRPB | Second caregiver believes in showing their love for me. | crpbi_caregiver15_y | Youth |
| Family/Home | Caregiver Support/Monitoring | CRPB | Second caregiver is easy to talk to. | crpbi_caregiver16_y | Youth |
| Family/Home | Caregiver Support/Monitoring | PM | How often do your parents/guardians know where you are? | parent_monitor_q1_y | Youth |
| Family/Home | Caregiver Support/Monitoring | PM | How often do your parents know who you are with when you are not at school and away from home? | parent_monitor_q2_y | Youth |
| Family/Home | Caregiver Support/Monitoring | PM | If you are at home when your parents or guardians are not, how often do you know how to get in touch with them? | parent_monitor_q3_y | Youth |
| Family/Home | Caregiver Support/Monitoring | PM | How often do you talk to your parent or guardian about your plans for the coming day, such as your plans about what will happen at school or what you are going to do with friends? | parent_monitor_q4_y | Youth |
| Family/Home | Caregiver Support/Monitoring | PM | In an average week, how many times do you and your parents/guardians, eat dinner together? | parent_monitor_q5_y | Youth |
| Family/Home | Caregiver Psychopathology | ASR | I scream or yell a lot | asr_q68_p | Caregiver |
| Family/Home | Caregiver Psychopathology | ASR | I use drugs (other than alcohol, nicotine) for nonmedical purposes | asr_q06_p | Caregiver |
| Family/Home | Caregiver Psychopathology | ASR | I drink too much alcohol or get drunk | asr_q90_p | Caregiver |
| Family/Home | Caregiver Psychopathology | ASR | I deliberately try to hurt or kill myself | asr_q18_p | Caregiver |
| Peer | Sexual Abuse | KSADS | A peer forced your child to do something sexually | ksads_ptsd_raw_769_p | Caregiver |
| Peer | Peer Victimization | KSADS | Does your child have any problems with bullying at school or in your neighborhood? | kbi_p_c_bully | Caregiver |

All items/measures were assessed at the baseline visit. Abbreviations of questionnaire names for Table S1 are as follows:

**KSADS**: Kiddie Schedule for Affective Disorders and Schizophrenia

**NSC**: Neighborhood Safety and Crime

**PDS**: Parent Demographics Survey

**FES**: Family Environment Scale

**CRPB**: Children's Report of Parental Behavior

**PM**: Parental Monitoring

**ASR**: Adult Self Report

**Table S2: Trajectory Features Used in Clustering Analysis^1^**

19 features calculated from longitudinal positive affect data

| Feature | Description | Interpretation |
| --- | --- | --- |
| Maximum | Highest positive affect value observed | Peak positive affect level |
| Range | Difference between maximum and minimum values | Overall variability magnitude |
| Mean Value | Average positive affect across all timepoints | Typical positive affect level |
| Standard Deviation | Variability in positive affect across timepoints | Within-person fluctuation |
| Intercept | Starting value from linear model fit | Baseline positive affect |
| Slope | Rate of linear change over time | Direction and magnitude of change |
| R² | Variance explained by linear trend | Linearity of trajectory |
| Curve Length | Total amount of change (sum of absolute changes) | Cumulative amount of fluctuation |
| Rate of Intersection with Mean | Frequency of crossing the individual mean | Stability vs. oscillation |
| Proportion Above Mean | Percentage of timepoints above individual mean | Tendency toward higher affect |
| Minimum 1st Derivative | Steepest decline in positive affect | Largest decrease |
| Maximum 1st Derivative | Steepest increase in positive affect | Largest increase |
| Mean 1st Derivative | Average rate of change | Overall direction of change |
| SD 1st Derivative | Variability in rate of change | Consistency of change |
| Minimum 2nd Derivative | Most negative acceleration (steepest deceleration) | Largest shift in trajectory |
| Maximum 2nd Derivative | Most positive acceleration | Largest shift in trajectory |
| Mean 2nd Derivative | Average acceleration/deceleration | Overall curvature direction |
| SD 2nd Derivative | Variability in acceleration patterns | Variability in curvature |
| Later/Early Change Ratio | Ratio of change in later vs. early period | Developmental timing of change |
| ^1^Features were calculated using the traj package in R | | |

**Table S3: Items Removed: Reduced Overlap 21-Item Scale^1^**

11 items removed to minimize positive affect overlap^1^

| **CBCL Item** | **Item Content** | **Overlapping PA Dimension(s)** |
| --- | --- | --- |
| **Anxious/Depressed** | | |
| **cbcl_q112_p** | Worries | Calm, At ease |
| **cbcl_q14_p** | Cries a lot | Positive emotion |
| **cbcl_q35_p** | Feels worthless or inferior | Confident |
| **cbcl_q45_p** | Nervous, highstrung, or tense | Calm, At ease |
| **cbcl_q50_p** | Too fearful or anxious | Calm, At ease, Confident |
| **cbcl_q52_p** | Feels too guilty | Confident, At ease |
| **cbcl_q71_p** | Self-conscious or easily embarrassed | Confident, At ease |
| **Withdrawn/Depressed** | | |
| **cbcl_q05_p** | There is very little he/she enjoys | Enthusiastic, Delighted, Interested |
| **cbcl_q102_p** | Underactive, slow moving, or lacks energy | Energetic |
| **cbcl_q103_p** | Unhappy, sad, or depressed | General positive affect |
| **cbcl_q75_p** | Too shy or timid | Confident |
| ^1^Original: 32 items (A/D: 13, W/D: 8, Somatic: 11); Reduced Overlap: 21 items (A/D: 7, W/D: 3, Somatic: 11) | | |

### **Figure S1. Distribution of ELA Z-score**

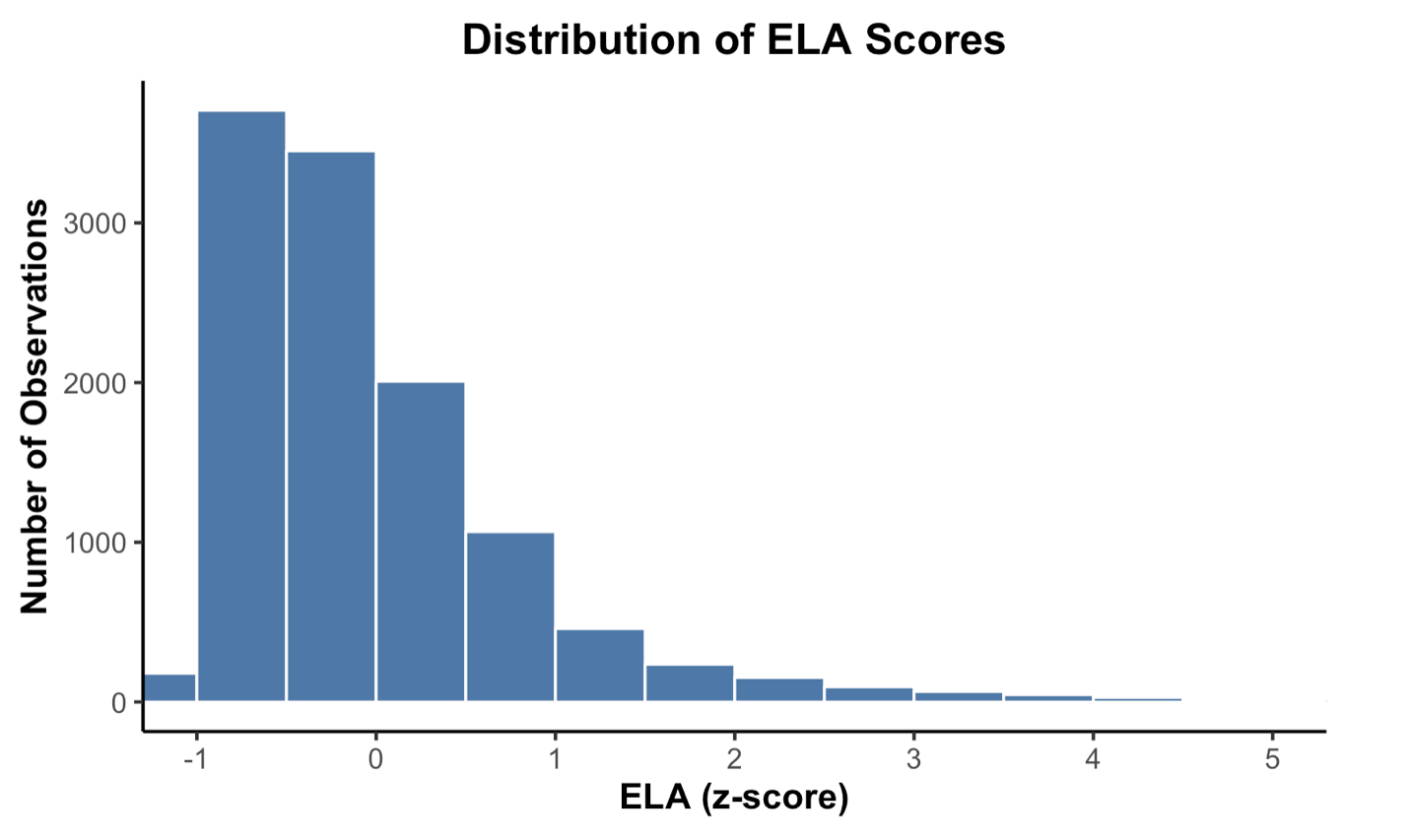

**Caption**: Distribution of Early Life Adversity (ELA) Composite Scores. Histogram showing the distribution of standardized ELA scores (z-scores). The distribution is right-skewed, with most participants experiencing relatively low levels of early adversity and fewer experiencing high levels. Of note, a small number of observations (n = 62) are not depicted on this histogram due to having values greater than 5. Of the total sample (N = 11,562), 4,230 participants (37%) had an ELA score greater than 0 while 7,335 participants (63.4%) had an ELA score lower than 0.

### **Figure S2. Mediation Using Alternative Formulations of Internalizing Subscales**

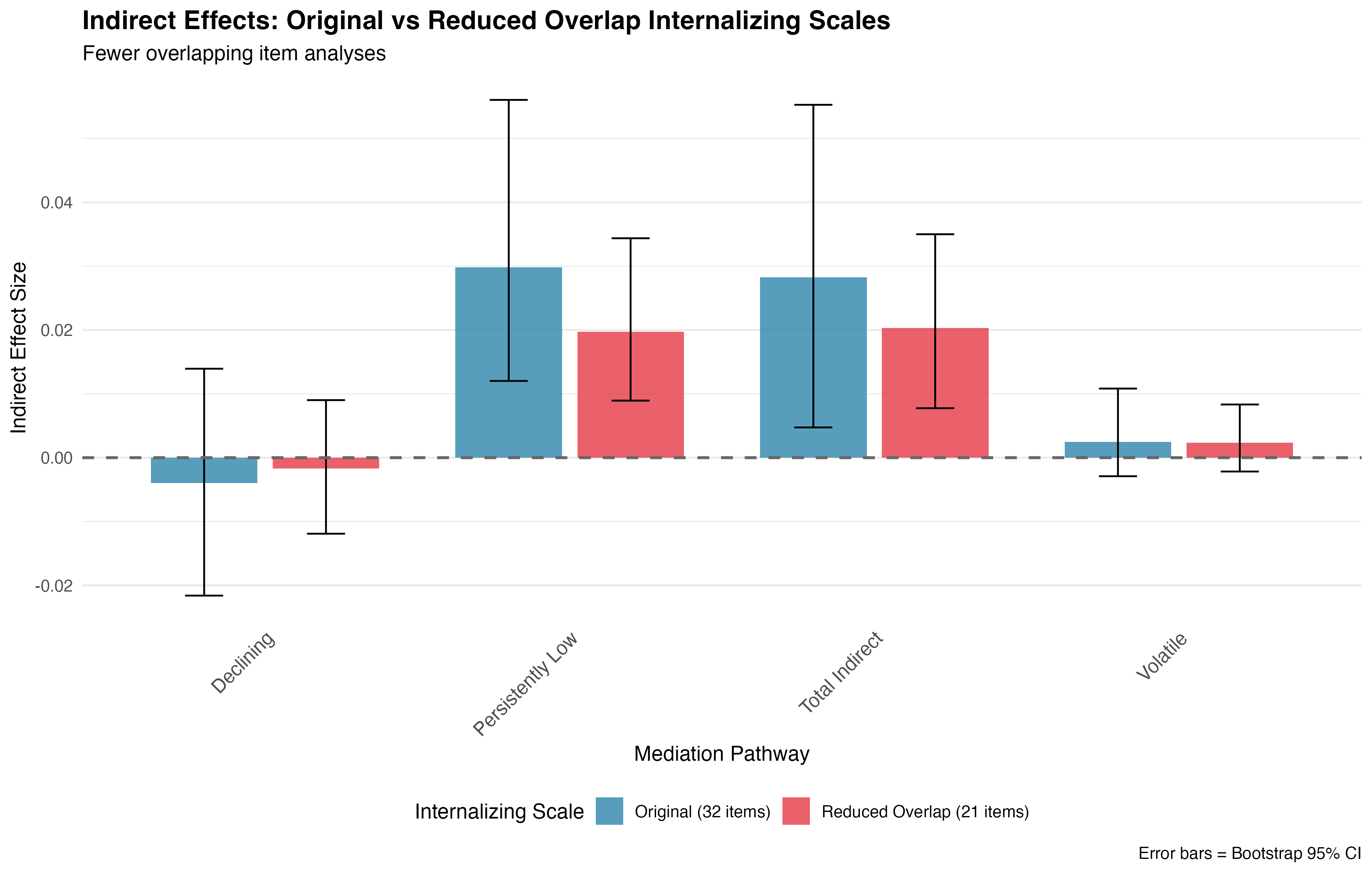

**Caption**: Indirect Effects Through PA Trajectories Using Original vs Reduced Overlap Internalizing Scales. Comparison of mediation pathways using the original 32-item internalizing scale versus a reduced overlap 21-item scale (r = .92). Both versions show significant total indirect effects and significant effects through the Persistently Low PA trajectory. Error bars represent bootstrap 95% confidence intervals.

**Figure S3. Missing Data Diagram.**

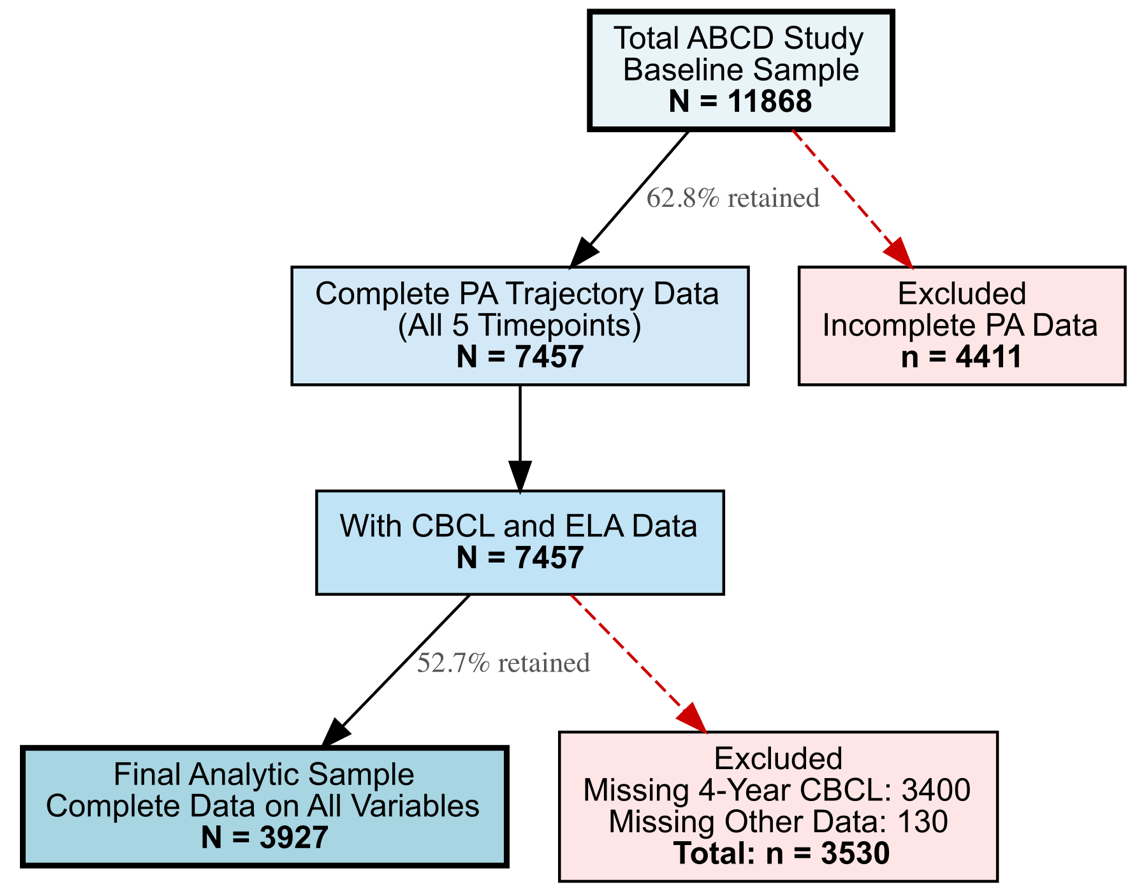

**Caption**: Participant Flow Diagram. Flow of participants from the ABCD Study baseline sample to the final analytic sample for mediation analyses. Primary attrition occurred due to incomplete positive affect (PA) trajectory data (37.2% excluded) and missing 4-year CBCL follow-up data (45.6% of those with PA trajectories excluded).
